# Mislabeled learning for psychiatric disorder detection

**DOI:** 10.1101/2022.08.11.22278675

**Authors:** Dongdong Li, Wenbin Liu, Henry Han

## Abstract

Mislabeled learning for high-dimensional data is essentially important in AI health and relevant fields but rarely investigated in machine learning. In this study, we address the challenge by proposing a novel mislabeled learning algorithm for high-dimensional data: psychiatric map diagnosis and applying it to solve a long-time bipolar disorder and schizophrenia misdiagnosis in psychiatry. The proposed algorithm converts each input high-dimensional SNP sample into a corresponding 2D characteristic image called a psychiatric map through feature self-organizing learning. It can automatically detect mislabeled observations and relabel them with the most likely ground truth before reproducible machine learning besides providing informative visualization for mislabeling detection. Our method attains more accurate and reproducible psychiatry diagnoses, besides discovering latent psychiatry subtypes not reported before. It works well for those datasets with a limited number of samples and achieves leading advantages over the deep learning peers. This study also presents new insight into the pathology of psychiatric disorders by constructing the devolution path of psychiatric states via relative entropy analysis that discloses latent internal transfer and devolution road maps between different psychiatric states. To the best of our knowledge, it is the first study to solve mislabeled learning for high-dimensional data and will inspire more future work in this field.

## 1 Introduction

With the surge of data amount and complexity in machine learning (ML), more and more mislabeled data challenges the existing machine learning models and AI methods. The mislabeled data generally refers to those data with incorrect information due to the artifacts of experimental design, system noise, or human mistakes. For example, some pathological images in AI health can be mislabeled probably due to doctors’ mistakes, the incidence of patient identification errors, or other unexpected issues. Almost all classic ML models would encounter mediocre or even poor performance under mislabeled data because they generally assume that data label information is correct by default. However, such an assumption may not match the existing data and reality, i.e., almost all data in AI practice can be mislabeled for various reasons, especially because data volume and complexity involved in learning have increased greatly in recent years. It was reported that mislabeled ratios in real world data generally can range from 8% to 38.5%, but some fields may have higher ratios [1,2]. For example, it was reported that more than a third of patients with severe psychiatric disorders such as schizophrenia and bipolar disorder were misdiagnosed (39.16%) in psychiatry [2,3].

The mislabeled data may corrupt the training procedure in AI and unavoidably produce misleading or wrong learning results by constructing biased or distorted decision functions. We call machine learning involved with mislabeled data as mislabeled learning. Investigating mislabeled learning has great practical significance, especially for various AI application domains. However, mislabeled learning is still not well investigated in the literature particularly for high-dimensional data for the following reasons.

First, most works focus on theoretical studies and deep learning models [2,4-8]. Some proposed techniques may not be completely automatic, and some level of human supervision may be needed to correct mislabeled samples [5,6]. In addition, it remains unknown whether the techniques developed for mislabeled learning under one ML model can apply to another one, because they may demonstrate different fault tolerance in learning.

Second, the proposed methods mostly focus on benchmark sample datasets (e.g., MNIST data) in deep learning. The datasets are not ‘truly mislabeled data’ by nature because they are artificially mislabeled with generated ‘incorrect labels’ with known ground truth. Compared to those ‘truly mislabeled data’ whose true labels are unknown, the former dataset can be more transparent and relatively easy to handle. As such, it remains unclear whether the proposed methods can apply to the mislabeled data from a specific application domain to another or not.

Third, more importantly, the proposed mislabeled learning methods generally may only apply to low-dimensional data rather than high-dimensional data, which is also widely used in all kinds of AI applications in health and medicine. The former is characterized by a large number of observations but a small number of variables. Most data in the existing mislabeled learning literature belong to low-dimensional data in which mislabeling occurs in the sample space with redundancy. On the other hand, the latter is characterized by a large number of variables (e.g., ∼O(10^4^)) but a small number of observations (e.g., ∼O(10^2^)). It suffers from the ‘curse of dimensionality in which mislabeling happens in a sparse sample space with limited entries, whereas the variable space has redundant information. Therefore, it can be hard to extend the existing mislabeled learning techniques developed for low-dimensional data to high-dimensional data. For instance, it can be infeasible to drop suspiciously mislabeled samples identified by the existing mislabeled learning techniques because of the very limited number of observations involved in learning [1,2].

As such, developing effective ML techniques to handle high-dimensional mislabeled data can be an urgent problem in applied AI and data science. High-dimensional mislabeled data brings more challenges in mislabeled learning than low-dimensional mislabeled data. Its sparse sample space contains mislabeling labels with unknown ground truth due to the ‘curse of dimensionality. To the best of our knowledge, there are almost no proposed methods to handle such data. On the other hand, since high-dimensional data is an essential component in AI health and precision medicine, efficient and effective high-dimensional mislabeled learning algorithms would not only bring positive impacts on the applied AI but also enrich the mislabeled learning techniques in ML.

In this study, we address mislabeled learning for high-dimensional data by solving a long-time bipolar disorder (BPD) and schizophrenia (SCZ) misdiagnosis in psychiatry. Schizophrenia (SCZ) and bipolar disorder (BPD) both are highly heritable, complex neuropsychiatric diseases that share some similar clinical symptoms [9]. SCZ is a chronic and severe mental health disorder characterized by hallucinations, delusions, and disorganized thinking. BPD is a chronic mental illness that causes dramatic shifts in a person’s mood, energy, and ability to think clearly. According to the National Alliance on Mental Illness (NAMI), it is reported that about 1% and 2.9% of Americans are diagnosed with SCZ and BPD respectively each year.

High-dimensional data, which is SNP data in our context, is widely employed to detect the molecular signatures of SCZ and BPD in various AI diagnoses [10-11]. However, it is essentially a high-dimensional mislabeled learning problem rooted in the misdiagnosis of SCZ and BPD. The label information of high-dimensional data is obtained from the clinical diagnoses that generally rely on the symptoms of patients rather than their real genetic differences. Since the symptoms of BPD and SCZ can be similar or even hard to distinguish for psychiatry doctors, a large portion of incorrect label information can appear due to the misdiagnosis, which also echoes their high misdiagnosis rates (e.g., >30%) in clinical practice [2-3]. The misdiagnosis between SCZ and BPD is a long-time challenging problem in psychiatry and presents a hurdle in AI-based diagnosis by generating mislabeled data in ML [11]. To some degree, solving the high-dimensional mislabeled learning problem is equivalent to overcoming the misdiagnosis between SCZ and BPD.

Recent bioinformatics studies show that SCZ and BPD may have different genetic differences. It was reported that there existed both unique and overlapping molecular signatures between SCZ and BPD [12]. Li *et al* suggested the different roles of DNA methylation in the pathogenesis of BPD and SCZ [13]. Ellis *et al* reported that the BPD and SCZ transcriptomes were not significantly correlated as originally expected [14]. Liu *et al* showed that SCZ and BPD shared common pathways and BPD could be a subtype of SCZ via manifold learning and pathway analysis [15]. Recent neuroimaging studies also reveal that SCZ and BPD patients can exhibit more characteristic patterns in brain imaging than normal people [16-18]. Since it is still premature to use both bioinformatics and neuroimaging results to correct the misdiagnosis between SCZ and BPD, the label information of the high-dimensional omics samples in AI diagnosis is inevitably noised and their ground truth remains unknown [15].

It can be more challenging to solve a high-dimensional mislabeled learning problem than a low-dimensional one because of the scarcity of samples and variable redundancy appearing in high-dimensional data. The small number of observations brings a serious issue in model selection. Those powerful deep learning techniques developed for mislabeled data generally need many samples to build an informative training space. Furthermore, the variable redundancy from a large number of features along with the mislabeled data may also bring difficulties in feature selection, i.e., many feature selection models need to know the ground truth, but it is not always available for mislabeled high-dimensional data, because the mislabeled information can inevitable by nature [15].

However, solving this problem will bring unpreceded impacts on AI health in which high-dimensional omics data is widely employed to conduct disease diagnosis and treatment. Technically, it will provide algorithmic support to advance high-dimensional mislabeled learning in AI health and interacting fields. Practically, it will shed new light on overcoming the long-time misdiagnosis of SCZ and BPD by providing more accurate diagnoses. The solving of misdiagnosis of SCZ and BPD not only brings a revolutionary breakthrough in psychiatry but also contributes to a deeper understanding of the pathogenesis of SCZ and BPD [19]. More importantly, it will inspire future mislabeled learning algorithm for those AI applications with only a limited number of observations by enriching machine learning

In this study, we propose a novel mislabeled learning algorithm: psychiatric map (*pMAP*) diagnosis for high-dimensional data by overcoming the long-time misdiagnosis problem between SCZ and BPD. Unlike other methods, the proposed algorithm converts each input high-dimensional SNP sample into a corresponding 2D characteristic image called a psychiatric map (*pMAP*) through an ML technique called feature self-organization maps (*fSOM)* learning. The mislabeled information correction is based on the downstream analysis and learning for the *pMAPs*. The proposed method, consisting of *fSOM* learning, DBSCAN clustering, and relabeled sample learning, can automatically identify and correct mislabeled samples in a rigorous, interpretable, and robust approach. Furthermore, it attains more accurate and reproducible BPD and SCZ psychiatry diagnoses besides discovering latent psychiatry subtypes not reported before.

As a special 2D characteristic image of each high-dimensional sample, the *pMAP* generated from *fSOM* learning is mathematically the prototype of the original 1D SNP sample. It summarizes the essential characteristics of each psychiatric sample as well as provides informative visualization. Theoretically, each *pMAP* is the low-dimensional nonlinear embedding of its corresponding high-dimensional sample in the subspace spanned by ‘condensed basis variables’ from *fSOM* learning. Each condensed basis variable is the low-dimensional embedding summarizing the essential characteristics of a set of original variables (SNPs). The total condensed basis variables summarize essential characteristics of all variables (SNPs).

The proposed *fSOM* learning is a customized self-organizing map for high-dimensional data to retrieve the prototype of each sample to unveil its ground truth characteristics. Unlike traditional self-organization map (SOM) that clusters input samples, *fSOM* seeks the nonlinear low-dimensional embedding *Z* ∈ ℝ^*k*×*n*^ of input high-dimensional data *X* ∈ ℝ^*p*×*n*^ with *n* observations across *p* variables (*p* ≫ *n, p* ≫ *k*) by implementing the map: *F*_*fsom*_: *X* → *Z* ∈ ℝ^*k*×*n*^ [20]. Each observation in the low-dimensional embedding *Z* has *k* condensed basis variables. Geometrically it is represented as a characteristic 2D image on the *fSOM* plane with 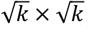 neurons.

Figure 1 illustrates the *pMAPs* of the control, BPD, and SCZ samples under *fSOM* using SNP data. It shows that those high-dimensional samples from the same groups share similar or the same 2D characteristic images (prototypes). On the other hand, the *pMAPs* of the control samples demonstrate clear differences from those of the BPD and SCZ samples. It suggests that there is no mislabeling occurrence between them. Such a result is consistent with the classification results in our previous studies, i.e., an SVM model can achieve nearly linearly separable performance to answer the query if a coming SNP sample is a control or diseased one [15].

**Fig 1.**
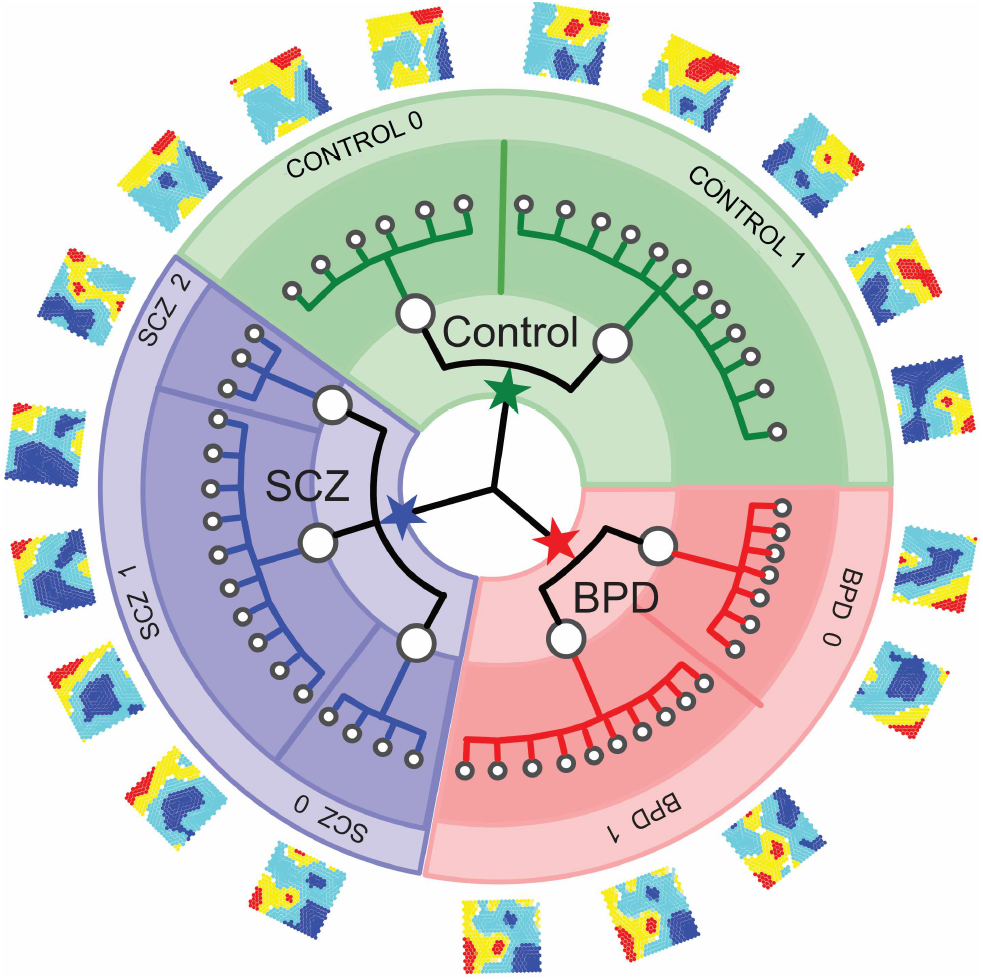
The psychiatric maps (*pMAPs*) of control, BPD, and SCZ samples obtained under *fSOM* learning by using SNP data. A *pMAP* is a 2D characteristic image of each high-dimensional psychiatry sample. *pMAPs* also unveil latent two control, two BPD, and three SCZ subtypes: *control_0/1, BPD_0/1*, and *SCZ_0/1/2*.

Besides the original three types of observations: control, BPD, and SCZ, the proposed algorithm employs *pMAPs* to unveil latent subtypes by identifying control 0/1 subtypes in the control group, and BPD 0/1 subtypes in the BPD group, and SCZ 0/1/2 in the SCZ group illustrated in Figure 1. It suggests that the algorithm is good at revealing latent information of high-dimensional data that contributes to identifying and correcting mislabeled samples well.

This study has the following contributions to mislabeled learning and psychiatry disorder diagnosis. First, it presents a novel mislabeled learning algorithm for high-dimensional data by enriching the existing mislabeled learning. To our knowledge, it is the first high-dimensional mislabeled learning algorithm. It includes novel techniques: *pMAP* generation using *fSOM* learning, *pMAP* clustering, and relabeled sample learning along with a data-driven feature selection algorithm nonnegative singular value approximation (nSVA) [21]. nSVA can conduct meaningful feature selection without the ground truth information for high-dimensional data. Unlike the existing mislabeled learning methods, our method can automatically detect mislabeled observations and relabel them with the most likely ground truth [1,5]. Furthermore, the proposed mislabeled learning algorithm does not require a large number of samples and works well for small-sample data with a limited number of samples. Therefore, it shows a good advantage in handling the sample scarcity issue in learning over almost all mislabeled learning peers [22].

Second, it provides a way to solve the long-time misdiagnosis of SCZ and BPD in psychiatry [15]. Compared to using bioinformatics approaches to identify possible driver genes, pathways, or other molecular signatures of SCZ and BPD, our AI-based approach is more reproducible, less expensive, and more applicable to accurate SCZ and BPD diagnoses [15]. Since the proposed method can discover latent subtypes by identifying previously not reported subtypes, it can provide valuable feedback to psychiatry doctors to examine and validate more latent SCZ/BPD subtypes [23].

Third, our study presents a novel visualization tool for high-dimensional data besides disclosing mislabeled samples. It maps each high-dimensional sample into a corresponding *pMAP*, a 2D characteristic image in the subspace spanned by ‘condensed key SNPs. To the best of our knowledge, it is the first work to convert a psychiatry disorder sample (SNP data) to a psychiatric characteristic image, i.e., *pMAP*, for the sake of knowledge discovery in mislabeled learning. Unlike existing mislabeled learning algorithms, the proposed *pMAP* diagnosis is a more explainable because it provides powerful sample visualization. On the other hand, it makes mislabeled learning techniques and SCZ and BPD misdiagnosis overcome more transparent and trustworthy. The knowledge-revealing *pMAPs* will assist doctors to understand the latent disease statuses/subtypes, achieve high-accuracy diagnoses, and enhance clinical decision-making.

Last but not least, we construct the devolution paths and internal transfers between different psychiatric states by conducting a novel relative entropy analysis to shed light on the pathology of psychiatric disorders. We also employ a new customized entropy analysis to explain the results of *pMAP* diagnosis in BPD and SCZ detection. We find that both BPD and SCZ samples tend to have lower entropies than the control samples. It may suggest that molecular patterns of the samples with psychiatric disorders should contain a ‘less random’ information pattern than the ordinary samples from SNP data analysis [10-11].

This paper is structured as follows. Section 2 introduces a high-dimensional mislabeled learning algorithm: *pMAP* diagnosis in details. Section 3 covers data and data preprocessing and section 4 presents the results of *pMAP* diagnosis from different perspectives. A novel devolution path model is developed in this section to explain the dynamics of psychiatric states by conducting relative entropy analysis for different *pMAPs*. Section 5 compares the proposed algorithm with the state-of-the-art ML and deep learning methods. Finally, we discuss the potential weakness and possible enhancements of our methods before concluding this study.

## 2 Psychiatric map (pMAP) diagnosis: a mislabeled-learning algorithm for high-dimensional data

A high-dimensional mislabeled learning problem can be described as follows. Given a training dataset 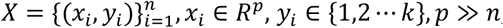, each sample (*x*_*i*_, *y*_*i*_) is assumed independent and identically distributed and the label information *y*_*i*_ can be corrupted and may not always reflect the ground truth of the corresponding sample *x*_*i*_. The goal of mislabeled learning is to seek a relabeling function 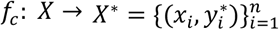 to map each sample *x*_*i*_ to its most likely ground truth 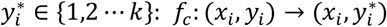, such that for an ML model *θ*, the following equation holds:

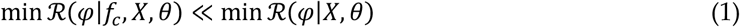

The empirical risks of the relabeled samples and the original samples under a loss function ***φ*** are defined as 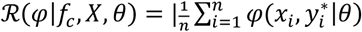 and 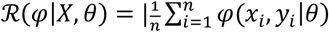 respectively.

It can be technically hard to attain a satisfactory learning performance by only applying existing ML models to high dimensional data with mislabeled information. It may only be possible when the percentage of corrupted labels is ignorable (e.g., 1%). However, SNP data in our context has over 30% mislabeled ratios. On the other hand, the existing mislabeled learning techniques may not be able to apply to the high-dimensional data because they are mainly developed for low-dimensional data. For example, adding a noise filtering in deep neural networks (DNN) may filter samples with corrupted labels, but it can’t apply to high-dimensional data because the filtering scheme can be too luxury to implement for a dataset only with limited number of samples [1,22]. Thus, it is necessary to seek an effective mislabeling correction technique to acts as the relabeling function *f*_*c*_ to tackle the challenge by retrieving the most likely ground truth for each sample.

It is desirable to extract the prototype of each observation to distinguish possibly mislabeled high-dimensional samples from an unsupervised learning approach before any possible label correction.

Compared to an original high-dimensional sample, the prototype is a small vector but contains the essential characteristics of the sample. The prototype should be able to identify itself without using any label information no matter whether it is true or false. The prototype should also provide visualization support so that its identity can be easily detected by comparing the 2D characteristic images of original samples. If samples A and B share similar or even the same prototype (2D characteristic image) but with different labels, it is easy to conclude the occurrence of mislabeling. Since it is also likely that many samples may share one or a few prototypes, the prototypes (2D characteristic images) can be an excellent discriminator to correct the mislabeled samples.

Which unsupervised learning approaches can generate the prototypes we need? Traditional dimension reduction techniques in unsupervised learning (e.g., PCA) may not be able to accomplish it because the low-dimensional embeddings from the methods are not representative enough to identify the original sample [24-25]. This is because of the built-in limitations of the methods. For example, PCA can only extract the global data characteristics of input data. The principal components, the bases of the PCA embedding space, are the linear combination of variables by maximizing the data variances globally [25]. As a result, the PCA embedding of a high-dimensional sample unavoidably misses the important local data characteristics that can be essential to identify two similar samples sharing some global similarities.

On the other hand, t-SNE, a classic manifold learning method, is good at extracting local data characteristics well rather than global ones [25-26]. It produces a low-dimensional embedding by minimizing the KL divergence between the Gaussian and student distributions modeling the sample similarities in the input and embedding space. The embedding is not strongly related to the variables of input data but the similarities of the input observation under a distance metric (e.g., Euclidean distance). As a result, the essential global data characteristics of high-dimensional data may not be caught in the t-SNE embedding. Besides, since t-SNE generates the embedding by solving a non-convex optimization problem, the embedding generation is unstable and not unique. Therefore, an unsupervised learning method to generate the prototype of input data should satisfy the following conditions.

1. It should generate the representative prototype (characteristic 2D image) of input high-dimensional data rather than only focusing on only global or local data characteristics.
2. It should produce a stable prototype strongly related to the variables of high-dimensional data by solving a convex optimization problem. The bases of the embedding space should consist of the condensed basis variables summarizing the essential characteristics of all variables. The learning procedure to generate the condensed basis variables should follow the similarities between the original variables rather than other externally imposed standards.
3. It can provide powerful visualization support for each prototype so that different prototypes can be compared visually for the sake of mislabeled sample identification.

To satisfy the conditions, we propose a novel feature self-organizing map (*fSOM)* learning to obtain prototypes (characteristic 2D images) of high-dimensional data. Unlike traditional SOM which dynamically looks for similarities between input samples, the proposed *fSOM* seeks to condense thousands of SNP features through self-organizing learning by seeking the similarities between SNPs to construct a low-dimensional embedding space spanned by the condensed SNPs summarizing the key characteristics of all SNPs. The *fSOM* is equivalent to solving a convex optimization via gradient learning to obtain prototypes to represent high-dimensional data without over-emphasizing on global or local data characteristics. Moreover, *fSOM* can provide powerful characteristic visualization for each prototype by visualizing the values of the corresponding embedding vector on an *fSOM* plane. In other words, *fSOM* maps each input high-dimensional sample to its 2D characteristic image. We name the prototype of each high-dimensional sample a psychiatric map (*pMAP*). In addition, we employ a purely data-driven method: nonnegative singular value approximation (nSVA) to select meaningful SNPs to handle the variable redundancy [15,21].

### 2.1 Psychiatric map diagnosis: a mislabeled learning algorithm for high-dimensional data

Figure 2 illustrates the flowchart of the proposed *pMAP* diagnosis. It consists of three major steps. 1) *pMAP* generation using *fSOM* learning. 2) *pMAP* DBSCAN clustering. 3) Relabeled sample learning besides other related post analysis.

**Fig 2.**
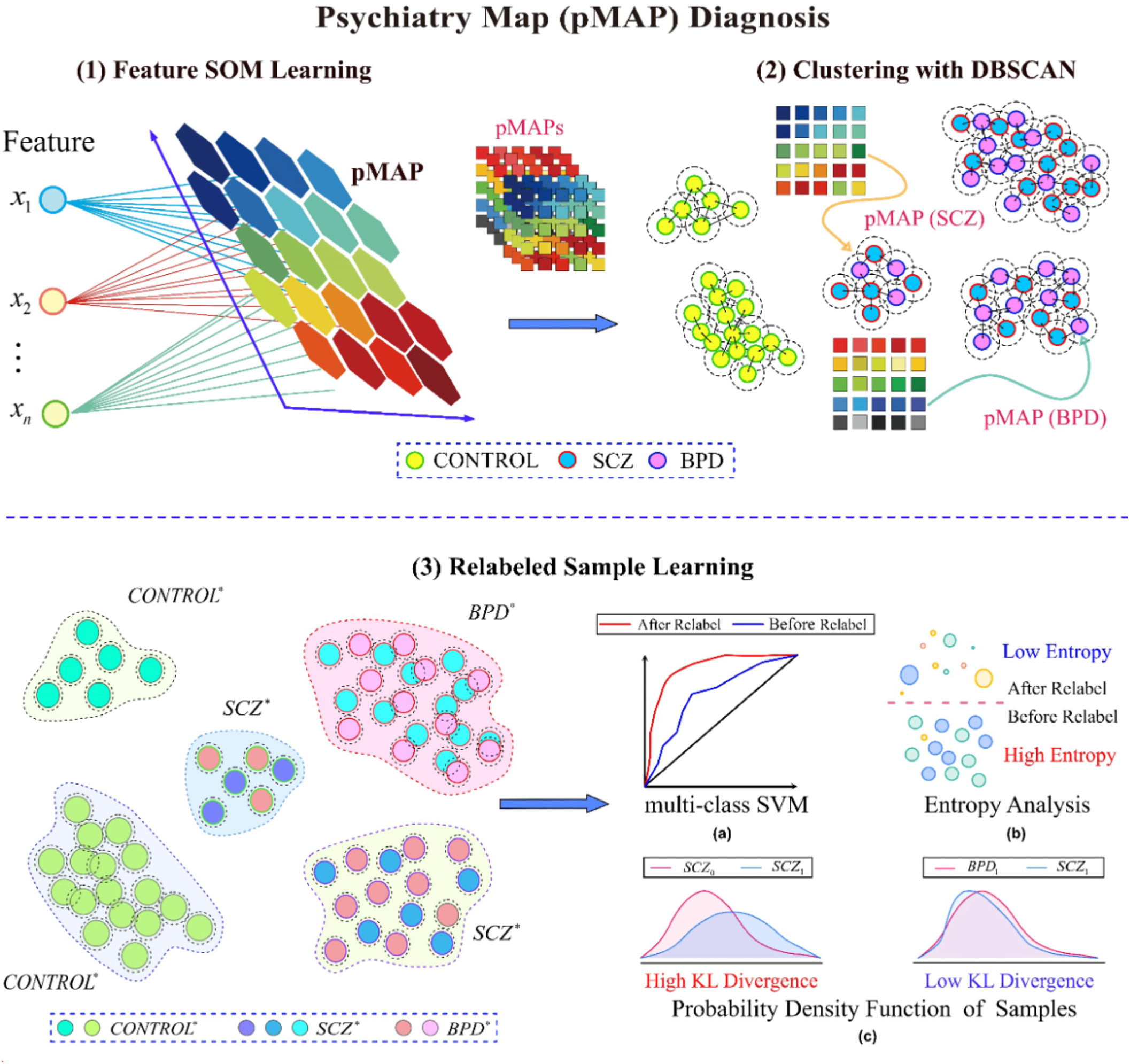
The flowchart of the proposed psychiatric map (*pMAP*) diagnosis. It employs feature self-organizing (*fSOM*) to generate a psychiatric map (*pMAP*), the prototype of each psychiatry observation which can be a schizophrenia (SCZ), a bipolar disorder (BPD), or a control sample. The prototypes then go through the DBSCAN clustering to seek the ground truth before relabeling each observation to its most likely ground truth [27-28]. Finally, a reproducible learning machine (e.g., multi-class SVM) is employed to conduct psychiatry diagnosis under the corrected labels besides different *pMAP*-based post analysis [29].

#### pMAP generation

employs *fSOM* learning to obtain the prototype of each SNP sample in the embedding space spanned by the *k* condensed basis SNPs: *S* = *span*(*s*_1_*s*_2…_*s*_*k*_ by implementing *F*_*fsom*_: *X* ∈ ℝ^*p*×*n*^→ *Z* ∈ ℝ^*k*×*n*^, *p* ≫ *n, p* ≫ *k*. For each condensed basis SNP, there exists a set of SNPs: 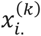 falling in a small enough neighbor of *s*_*k*_ with radius 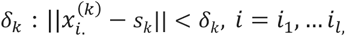. Geometrically, a *pMAP* is the characteristic image of an original high-dimensional sample. Mathematically a *pMAP* is the reference vector in ℝ^*k*^, *k* ≪ *n*, of a high-dimensional sample on the 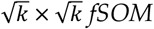 plane. In addition, we find that the pairwise distance between two *pMAPs* in the embedding space induced by *fSOM* learning will be less than their original high-dimensional samples in the input space | |*F*_*fsom*_(*x*_.*i*_) – *F*_*fsom*_(*x*_.*j*_)|| < ||*x*_.*i*_ − *x*_.*j*_|| because of the dimensionalities of the condensed basis SNP under self-organizing map for features.

The *pMAPs* will demonstrate good sensitivity for mislabeled sample identification. Since each *pMAP* is the prototype of a psychiatry sample, it is natural that the samples from the same psychiatry class will share similar or the same prototype, but the mislabeled samples will be those labeled with the same psychiatry type but with very different *pMAPs* or those with similar or the same *pMAPs* but with different labels.

#### *pMAP* DBSCAN clustering

The *pMAP* diagnosis employs a robust density clustering algorithm: DBSCAN (density-based spatial clustering of applications with noise) to cluster the *pMAPs* of input samples in the *fSOM* embedding space to correct possible mislabeled samples. The clustering of the 2D characteristic images of input data answers the following query: which observations are similar enough to fall in the same cluster?

It is noted that the ground truth is unknown because of the misdiagnosis between SCZ and BPD. There are two reasons for us to select the density-based clustering method DBSCAN rather than the popular K-means. The first is that K-means would require input data is convex. But we cannot guarantee the *pMAPs* will satisfy it. Instead, we find that *pMAPs* can have concaved shapes. The second is that K-means would limit the possible new subgroup detection somewhat. K-means may look like a good candidate for *pMAP* clustering because we already know the prior three groups: control, SCZ, and BPD. However, the *pMAP* generation stage can bring ‘new’ prototypes that are not limited to the original ones. Instead, the *pMAPs* unveil new knowledge, i.e., the control, SCZ, and BPD groups all have their different subtypes. Thus, we do need a clustering algorithm like DBSCAN that can automatically identify the number of clusters for input *pMAPs* for the sake of deep knowledge discovery [27].

The following proposition states that the two psychiatry samples will share the same ground truth if their *pMAPs* fall in the same cluster and when both are classified as the core points. A core point in DBSCAN is the point whose neighborhood with an enough number of points to form a ‘dense region’ [27]. Given a set of observations *X* = {*x*_1,_*x*_2,_ ⋯ *x*_*n*_}, a neighbor radius *ε*, and an integer *minpts* > 0, *x* ∈ *X* is a core point if and only if |{*x*^2^: |*x* − *x*^2^| < *ε*}| ≥ *minpts*, in which *minpts* is the minimum number of points to form a ‘dense region’ in DBSCAN. More details about DBSCAN can be found in Section 2.3.

##### Proposition 1.

Given any two high-dimensional samples *x*_.*i*_ and *x*_.*j*_ if their *pMAPs F*_*fsom*_(*x*_.*i*_) *and F*_*fsom*_(*x*_.*j*_) both fall in the same cluster under DBSCAN and both of them are the core points under DBSCAN, then Pr {*y*_*i*\*_ ≠ *y*_*j*\*_| |*F*_*fsom*_(*x*_.*i*_) − *F*_*fsom*_(*x*_.*i*_) |< *ε*} → 0, in which *y*_*i*\*_ and *y*_*j*\*_ are the ground truth labels of *x*_.*i*_ and *x*_.*j*_ respectively, and *ε* is the neighborhood radius in DBSCAN.

#### Relabeled sample learning

We relabel the original samples and their *pMAPs* according to the DBSCAN clustering result before employing a reproducible ML model to conduct psychiatry diagnosis. Given two psychiatric high-dimensional samples *x*_.*i*_ and *x*_.*j*_ and the DBSCAN clustering indices *z*_*i*,_*z*_*j*_ of their *pMAPs*: *z*_*i*=_*F*_*fsom*_(*x*_.*i*_), *z*_*j*=_*F*_*fsom*_(*x*_.*j*_): *z*_*i*,_*z*_*j*←_*dbscan*(*z*_.*i*_, *z*_.*j*_), then the labels of *z*_.*i*_, *z*_.*j*_ are updated by their clustering indices *l*_*i*_ ← *z*_*i*_ and *l*_*j*_ ← *z*_*j*_ respectively. So do the labels of *x*_.*i*_ and *x*_.*j*_.

Although we have found the previously unknown subtypes for each psychiatric group in the *pMAP* generation, we still label those from the same group as one type rather than dividing them into different types for the sake of learning and peer comparisons. For examples, two different control subtypes are found in the *pMAP* generation and corresponding two clusters are founded in the following DBSCAN clustering. We still label the *pMAPs* and their original samples as the original control type according to the original label of the core points rather than assign a new label. The following proposition 2 describes the relabeling rule under this situation.

##### Proposition 2

Given *pMAPs z*_.*i*_ and *z*_.*j*_ with different DBSCAN clustering indices *z*_*i*,_ ≠ *z*_*j*_, if their original samples *x*_.*i*_ and *x*_.*j*_ share the same label *y*_*i*_ and the label *y*_*i*_ shared by the original samples of the majority of the core points of the clusters *z*_*i*_ and *z*_*j*_, then the new labels of *z*_*i*_ and *z*_*j*_ will be assigned as *l*_*i*_ = *l*_*j*_ ← *y*_*i*_.

It may be possible theoretically to label those *pMAPs* and their original samples with the new subtypes unveiled in clustering. However, it would potentially create a learning risk because it is likely that some subtypes have only a few samples. The scarcity of samples in a certain type will generate inaccurate or even biased learning results under the situation [25, 40].

#### Reproducible learning model selection

After the relabeling procedure, an ML model is used to conduct diagnosis for the relabeled psychiatric maps. Theoretically, any ML models can be employed, but we prefer the ML model to satisfy the following standards for high-performance diagnosis.

First, it should have good reproducibility so that the psychiatry diagnosis would not change from run to run. Good reproducibility is important in clinical practice to decrease false positive rates and build customers’ trustworthiness. In other words, we should avoid those ensemble learning methods such as random forests or deep learning models because their results may lack good reproducibility for their built-in randomness in learning, though their learning performance can be good [30].

Second, it should have a built-in advantage to handle high-dimensional data with a small number of samples. In other words, its complexity should not increase much for high-dimensional data. The deep learning models generally need a large amount of data in training to build sophisticated prediction functions, but they are not good for high-dimensional data. This is because almost all deep learning models suffer from the data scarcity problem, i.e., deep learning may have poor performance for datasets with only a limited number of observations [22].

In this study, we employ multi-class support vector machines (multi-class SVM) because it satisfies the standards well. SVM demonstrates good reproducibility because it is equivalent to solving a deterministic nonlinear programming problem with the least randomness compared to the ensemble learning and deep learning methods. Furthermore, SVM is good at handling high-dimensional data because kernel matrix computing will be efficient for its small size. It contributes to speeding up the whole SVM learning by avoiding possible computing overhead. The one-versus-one (‘*ovo*’) scheme is employed to extend relevant binary SVM to corresponding multiclass diagnoses. It decomposes the *k*-class classification problem as the combination of *k(k-1)/2* binary SVM classification problems. The reason for using the ‘*ovo*’ rather than peer one-versus-rest (*‘ovr’*) multi-class handling scheme is that the latter may face an imbalanced learning issue [31]. We briefly describe the binary SVM model as follows.

A binary SVM model constructs an optimal hyperplane *y* = *w*^*T*^*z* + *b* to separate two groups of *pMAPs* of psychiatry SNP data under the training data 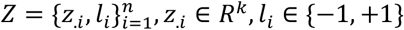 to by solving a quadratic programming problem:

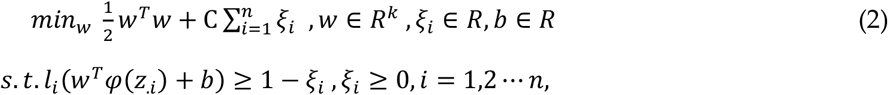

It is noted that *z*_.*i*_ ∈ *R*^*k*^ is a *pMAP* of the original SNP sample *x*_*i*_ and *l*_*i*_ is the label from the relabeling procedure. The normal vector *w* is in *R*^*k*^, in which *k* is the number of neuros on the *fSOM* plane, C ∈ *R*^+^ is the regularization parameter, and *φ*(·) is an implicit feature function mapping input data to the high-dimensional feature space for evaluation using kernel tricks. The Gaussian kernel 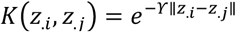, ϒ > 0 is mainly employed to model nonlinear relationships. The kernel parameter ϒ is tuned under the grid search technique for the sake of effective diagnoses [32].

The following Algorithm 1 summarizes the proposed psychiatric maps (*pMAP*) diagnosis. The complexity of the proposed algorithm complexity is *O*(*npk* + *plogp*) + *O*(*θ*), where *n* and *p* are the number of observations, features of input data and *k* is the number of neurons on the *fSOM* plane. *O*(*θ*) is the complexity of multi-class SVM used in psychiatric map diagnosis, i.e., *Ο*(*θk* = *Ο*(*nk*^2^).

##### Algorithm 1

**Psychiatric map (pMAP) diagnosis**

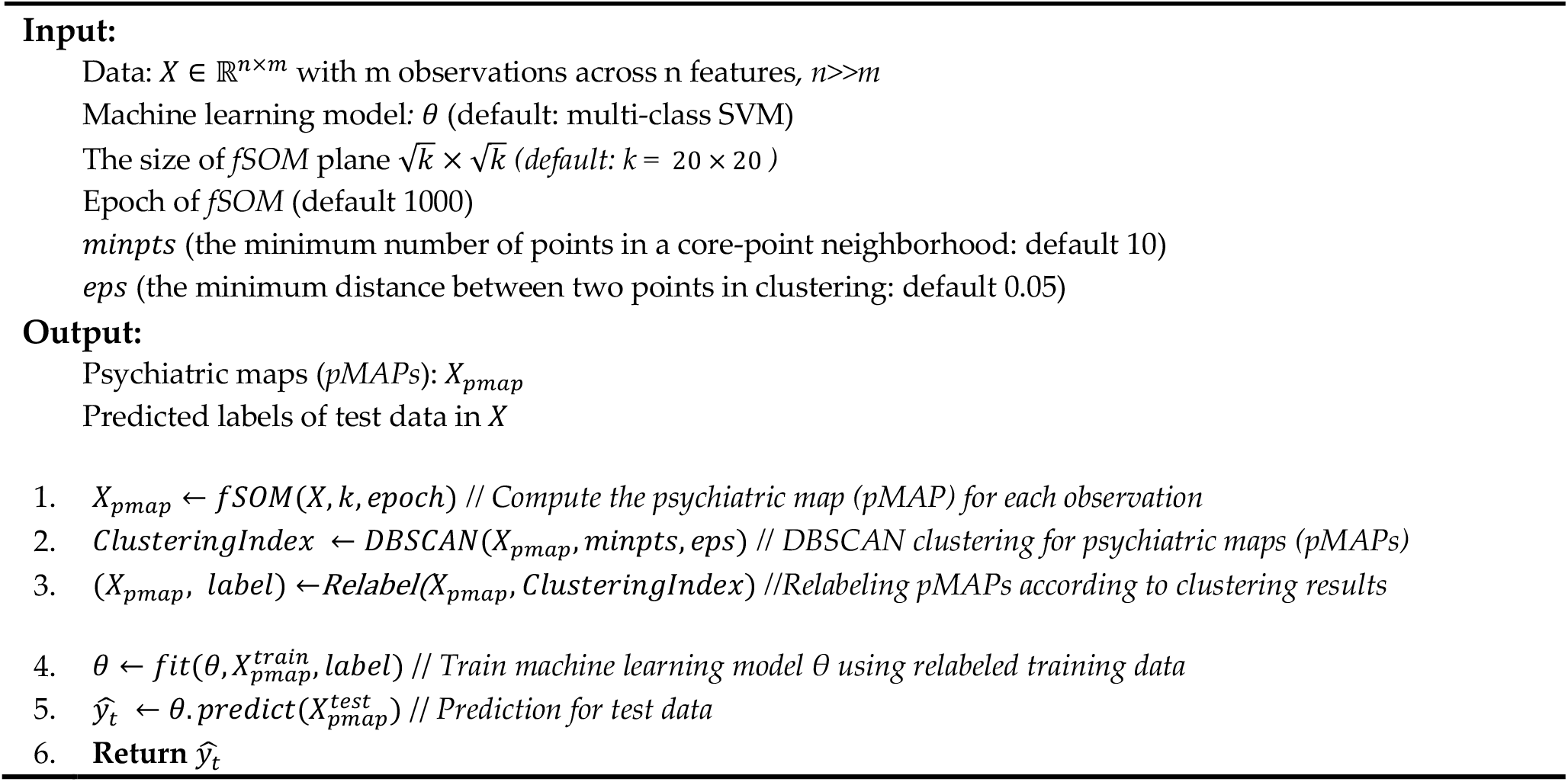

In the post-analysis of the *pMAP* diagnosis, we propose novel entropy and relative entropy (KL divergence) analysis to quantify relabeled psychiatric samples and investigate possible devolutionary relationships between them. Traditionally, it is almost impossible to compute the metrics for a group of psychiatric samples because they are not defined for a matrix originally. With the help of the *pMAPs*, we innovatively calculate them and examine possible devolution paths of psychiatric states and find that the psychiatric disorder samples may have more special SNP patterns than those of the normal ones. Please see more relevant details in Section 4.

### 2.2 Feature self-organizing map (*fSOM*) learning

Unlike traditional SOM, feature self-organizing map (*fSOM)* assumes input data is high-dimensional data and seeks the prototype of input data by conducting self-organizing learning for features rather than sample clustering.

*fSOM* consists of an input high-dimensional dataset, in which each row is a feature, and each column is an observation, *X* ∈ ℝ^*p*×*n*^, *p* ≫*n*, an *fSOM* plane 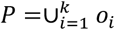, a two-dimensional lattice consisting of 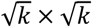 *(e*.*g*., *k=*400) neurons, and a loss function *ℒ* to define the embedding error in obtaining the prototype data, i.e., *fSOM* = (*X, P, ℒ*). Each neuron *o*_*i*_ ∈ *P* is associated with an embedding vector *z*_*i*_ ∈ ℝ^*k*^. Given input data with *n* samples across *p* features, *fSOM* produces its prototype *Z* by completing the nonlinear dimension reduction mapping: *F*_*fsom*_: *X* ∈ ℝ^*p*×,^ → *X* ∈ℝ^)×,^, *p* ≫*n, p* ≫*k* in a low-dimensional embedding space, in which *Z* = [*z*_.1,_*z*_.2_ ⋯ *z*_.*n*_] *is the* the extracted prototype data of input data *X*. The each column of the embedding data *Z* represents a corresponding prototype of an input sample. The embedding space is spanned by the condensed basis variables: *s*_1_, *s*_2_ ⋯ *s*_*k*_ in ℝ,, which are technically the row vectors of the embedding data: *s*_*i*_∼*z*_*i*._, *i* = 1,2 ⋯ *k*, in which *z*_*i*._ represents the *i*^*th*:^ row of the embedding data *Z*.

Each condensed basis variable 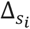 is the characteristic variable of a set of most similar variables: 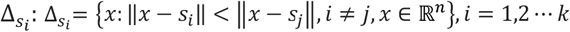. As a small set of characteristic variables, the condensed basis variables summarize the original high-dimensional variable space according to different characteristics of variables.

#### The *fSOM* learning: competition, cooperation, and adjusting

The *fSOM* learning consists of a loop of three stages: competition, cooperation, and adjusting. In the competition stage, given a feature *x* ∈ *X, fSOM* queries all neurons *o*_*i*_ ∈ *P, i* = 1,2 ⋯ *k* to find a neuron candidate *o*_*j*\*_ whose associated embedding vector *z*_*j*\*._ is nearest to *x* according to a distance metric (e.g., Euclidean): 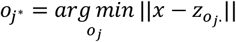. The candidate *o*_*j*\*_ will be the winning neuron in the competition stage and called the best match unit (BMU) for *x*.

In the cooperation stage, the embedding vectors associated with the neurons in the neighborhood of the winning neuron *o*_*j*\*_ is updated to make them more and more ‘similar’ to the feature *x* by following the iteration scheme:

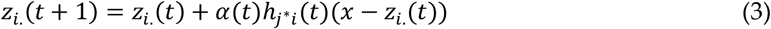

where *α*(*t*)*k* ∈ (0,1) is the learning rate at time *t*. The neighborhood function *h*_*j*\**i*_ models the proximity degree of the neurons in the neighborhood with respect to the winner. The function is chosen as a Gaussian kernel 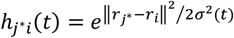 in this study for its fast convergence, in which *r*_*j*\*_ and *r*_*i*_ are the topological locations of the winning unit *o*_*j*\*_ and *i*^*th*^ neuron *o*_*i*_ on the *fSOM* plane, and the parameter *σ* denotes the neighborhood radius.

In the adjusting stage, the learning rate is adjusted, and the neighborhood radius decreases with respect to time exponentially to localize the iteration. That is, 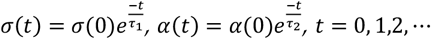. The *α*(0) and *σ*(0) are the initial learning rate and neighborhood size respectively and τ_1_ and τ_2_ are pre-selected time constants. The three stages are repeated until the embedding vector matrix *Z* converges: ‖*Z*(*t* + 1) − *Z*(*t*)‖ < η (10^−2^), or an expected epoch reached.

The *fSOM* learning is equivalent to a gradient-based optimization with the loss function 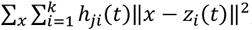, defined in a neighborhood centered in the winning neuron *j*, where the distance measure can be any specified distance metric. Thus, the iteration scheme is equivalent to finding the optimal embedding vector of a neuron to minimize the loss function through gradient optimization problem:

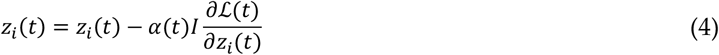

where *I* is an identity matrix and *α*(*t*)*I* is the learning rate matrix. The computing complexity of one epoch of training to minimize the loss function is *O*(*npk*). It is theoretically possible to a take second-order Newton method to accelerate the learning, but its effectiveness may rely on the initial point selection and the size of the neighborhood.

Note that the prototype data *Z* = [*z*_.1,_*z*_.2_ ⋯ *z*_.*n*_] can serve as a better embedding to represent the original data than those PCA, t-SNE, or UMAP embeddings because it is built by partitioning the variable spaces into different condensed basis variables through self-organizing learning [33]. It avoids possible information loss in PCA, t-SNE, or UMAP for each sample in embedding by over-emphasizing global or local data characteristics as well as overcomes the instability of t-SNE/UMAP embeddings., The prototypes can provide powerful visualization to distinguish possible mislabeled samples. Therefore, each prototype can be a valuable information source for overcoming corrupted or noisy labels in mislabeled learning. We have the following results about the nonlinear embedding obtained from *fSOM*.

##### Proposition 3.

Given high-dimensional samples *x*_.*i*_, *x*_.*i ′*_ and *x*_.*j*_ with corresponding labels *y*_*i*_, *y*_*j*,_ and *y*_*j*,_ *y*_*i*_ ≠ *y*_*j*,_ in which the ground truth label of *x*_.*i ′*_ is *y*_*i*_, then *z*_.*i*_ and *z*_.*i ′*_ will be more likely to fall in the same cluster than *z*_.*i*_ and *z*_.*j*_ under a clustering algorithm Π, in which *z*_.*i*_, *z*_.*i ′*_ and *z*_.*j*_ are corresponding embeddings of *x*_.*i*_, *x*_.*i ′*_ and *x*_.*j*_ under *fSOM*.

### 2.3 Density-based spatial clustering of applications with noise (DBSCAN)

DBSCAN is a density-based clustering algorithm that handles arbitrary-shaped clusters with noise [27-28]. DBSCAN classifies points as core, reachable, and outliers (noise). A core point simply refers to a point whose neighborhood has enough points under a radius *ε*. A reachable point is a point that can be reached by one or a sequence of core points and an outlier is an unreachable point, i.e., noise. In our context, it will be a transaction with exceptional trading behaviors that are potential to be trading markers. The core points form clusters because of their high densities, the reachable points form the edge of clusters, and the outliers stand out as noise in clustering.

The primary idea of DBSCAN can be described briefly as follows. Given a point to be clustered, DBSCAN retrieves its ε-neighborhood. If the neighborhood size is ≥ the minimum number of points (*minpts*) required to form a ‘dense region’, i.e., a region with an enough number of close points, the neighbor will be initialized as a cluster and the point is marked as a core point. Otherwise, the point is marked as an outlier. If the point is a reachable point for a cluster, its ε-neighborhood will be marked as a part of that cluster. All points in the ε-neighborhood will be added to the cluster until the density condition is satisfied. This procedure continues until all clusters and outliers are identified. The average running time complexity of DBSCAN is Ο(*nlogn*) if a meaningful neighborhood radius ε is selected, though the worse time complexity is Ο(*n*^2^).

## 3 Data and preprocessing

The original SNP data (GSE71443) downloaded from the NCBI GEO database includes 74 control, 65 bipolar disorder (BPD), and 64 schizophrenia (SCZ) subjects [35-36]. To obtain the significant differentially expressed SNP loci, we first filter those SNPs with missing annotations, not on autosomes or sex chromosomes, or diverged from Hardy-Weinberg equilibrium (HWE) with *p*_*value* < 10^−10^ [35]. We still have a total of 627,693 SNPs left after the initial filtering. We further screen statistically significant SNPs using ANOVA with *FDR* < 0.01 and remove those SNPs in the linkage disequilibrium (LD) using *R*^2^ > 0.25 [35]. Finally, we obtain a dataset with 5,843 SNPs across 74 control, 65 BPD, and 64 SCZ samples. Although it is theoretical to conduct *fSOM* for the preprocessed data, it may be desirable to seek more important features for the following unsupervised learning and *pMAP* diagnosis besides lowering the complexities caused by the high dimensionality.

As we mentioned before, the variable redundancy in mislabeled high-dimensional can bring difficulties in feature selection because many feature selection models need to know the ground truth. We seek the most critial features from the preprocessed SNP data by employing nonnegative singular value approximation (nSVA), an effective data-driven feature selection algorithm proposed by Han for high-dimensional data that does not need ground truth information [21]. Unlike the traditional model-driven methods that generally assume SNP data subject to a specific probability distribution, nSVA feature selection does not need the prior probability distribution besides ground truth information. It exploits the nonnegativity of input SNP data and ranks the importance of features according to its projection onto the first singular vector direction. Our previous work shows that it can identify meaningful feature selection for high-accuracy downstream analysis such as classification and pathway analysis [15,21].

## 4 Results

The *pMAP* generation *from fSOM* learning not only unveils new knowledge by discovering the latent subtypes of control, BPD, and SCZ, but also obtains data distributions that cannot be computed from input high-dimensional data. It somewhat provides more information sources for mislabeled sample correction. Unlike the original assumptions that samples are partitioned as control, BPD, and SCZ groups, the *pMAPs* discover that there are two subtypes for control, two subtypes for BPD, and three subtypes for SCZ. It suggests that *pMAPs* provide a meaningful knowledge discovery process to disclose intrinsic data characteristics for mislabeled data correction. On the other hand, *pMAPs* also provide a powerful visualization technique to discover different new subtype information for the original samples to identify mislabeled samples visually.

### Mislabeled sample visualization

Figure 3 illustrates the *pMAPs* of the three types of samples as well as their new subtypes discovered in *fSOM* learning. Figure 3 (a) shows that control has two types of psychiatric maps named ‘*control*’ type 0 and 1, both of which have blue regions, in which the embedding vectors have small values, on the right boundary of the *fSOM* plane. The *pMAPs* of the controls demonstrate quite clear differences from those of the BPD and SCZ samples. This concurs with our previous results that general machine learning can achieve 98% diagnostic accuracy between control and BPD/SCZ, which is approximately a linearly separable problem [15]. Figure 3 (b) shows that the SCZ samples consist of three different hidden types of *pMAPs*, named SCZ 0, 1, and 2 respectively. The *pMAPs* of SCZ 0 (‘type 0’), SCZ 2 (‘type 2), and BPD 0 (‘type 0’) share very similar patterns indicating they are highly likely to be mislabeled ones. For example, the *pMAPs* of SCZ 0 and BPD 0 share very similar patterns, but they are labeled as different types. Therefore, those samples (e.g., SCZ 0 and BPD 0) with similar *pMAPs* but different labels are highly likely to be mislabeled ones from their *pMAPs*.

**Fig 3.**
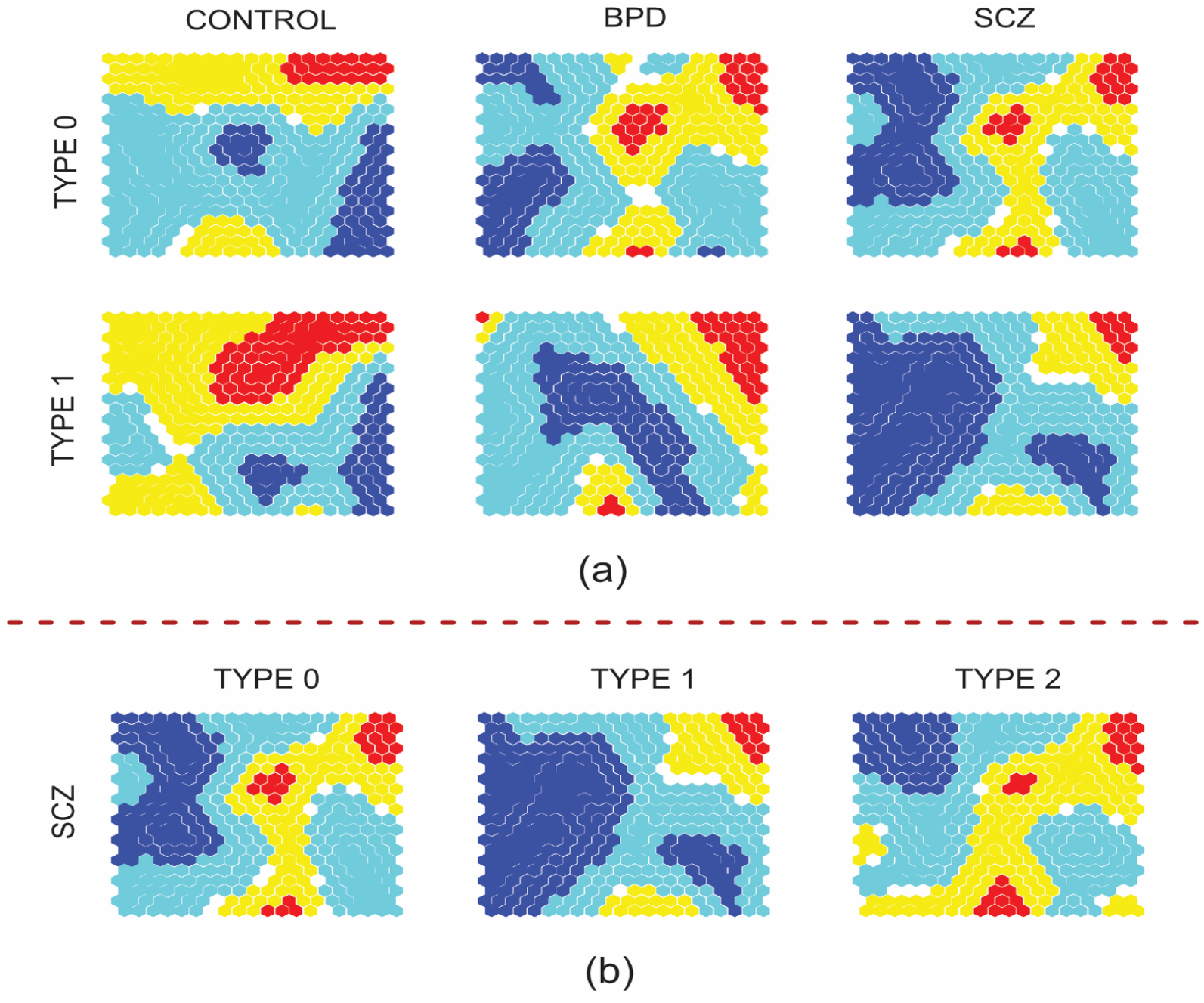
The *pMAPs* of control, BPD, and SCZ generated from *fSOM* learning. (a) shows the type 0 and 1 *pMAPs* of control, BPD, and SCZ. (b) illustrates the *pMAPs* of all the three discovered SCZ subtypes. The *pMAPs* of controls demonstrate obvious differences from those of the BPD and SCZ. The *pMAPs* of SCZ 0 (‘type 0’), SCZ 2 (‘type 2), and BPD 0 (‘type 0’) share very similar patterns indicating they are highly potentially mislabeled ones.

### Psychiatric type probability density function estimation

The *pMAPs* also provide a powerful way to estimate the probability density function (*p*.*d*.*f*) for each psychiatric type, which is almost impossible for input high-dimensional data even if it does not have noisy labels. Such an estimation contributes to detecting mislabeled samples in a more rigorous way. We apply Gaussian kernel density estimation to the *pMAP* of each sample, which is further reshaped as a 1-D vector, to estimate the probability density functions of different types of samples [37]. To the best of knowledge, it is the first work in estimating the *p*.*d*.*f*. of high-dimensional mislabeled data.

Figure 4 compares probability the *p*.*d*.*f*.s of the subtypes of the control, BPD, and SCZ samples, where the horizontal direction represents the gene expression levels. The subfigures from (a) to (c) summarize the *p*.*d*.*f*.s of the subtypes in each group and indicate their obvious differences, where the *p*.*d*.*f*.s of the two control subtypes have very different skewness. For example, the skewness and kurtosis values of ‘control 0’ are 0.9946 and -0.3445, but those of ‘control 1’ are 0.3517 and -1.3085. The subfigures from (d) to (i) pairwisely compare the *p*.*d*.*f*.s of the subtypes of the BPD and SCZ groups. It also strongly suggests the similarity between the subtypes BPD 0, SCZ 0, and SCZ 2, which may indicate the occurrence of mislabeled types.

**Fig 4.**
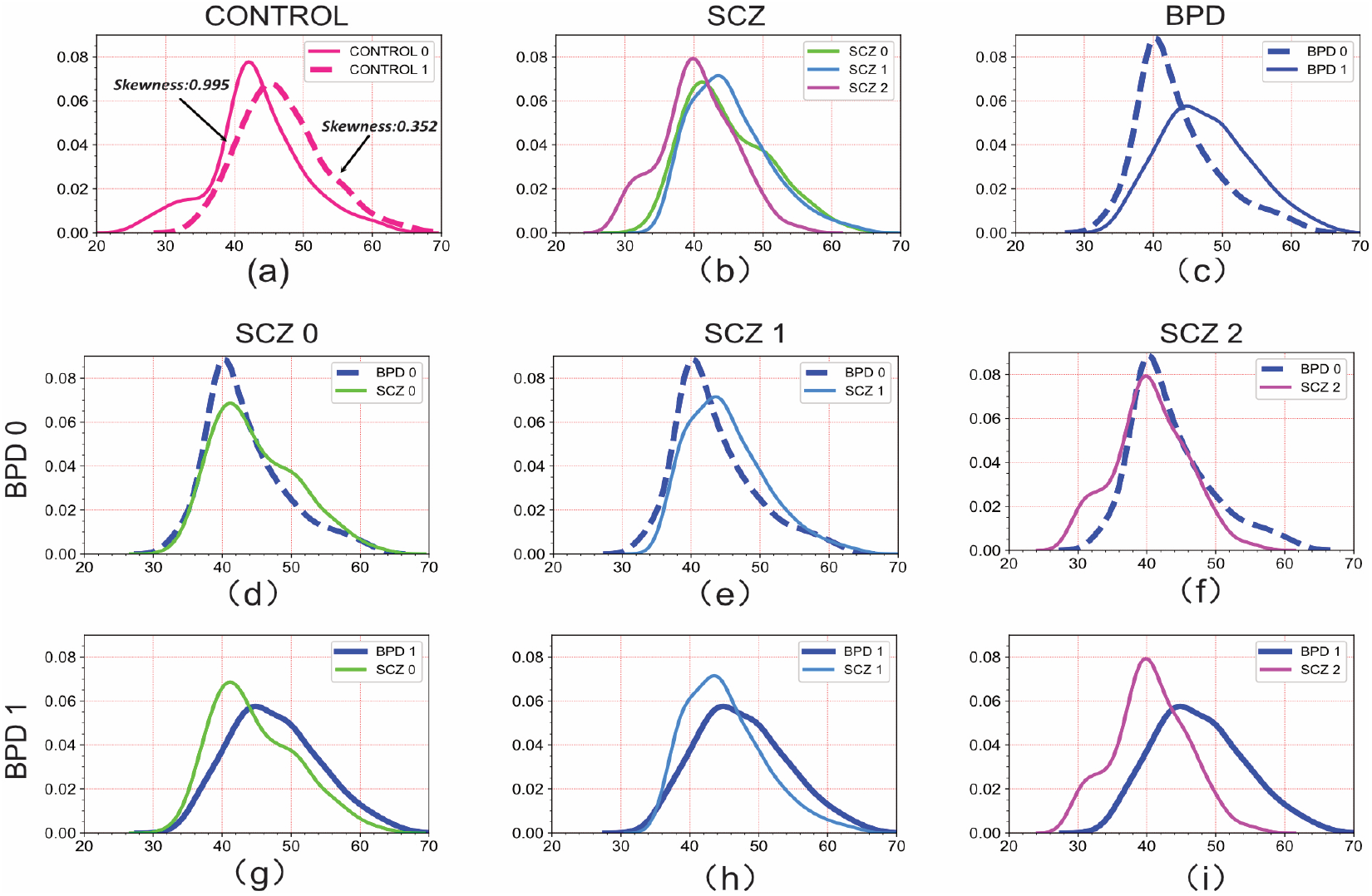
The *p*.*d*.*f*.s of the different subtypes of the control, BPD, and SCZ samples. The subplots (a) to (c) illustrate the *p*.*d*.*f*.s of the subtypes of the three groups and suggest quite good differences between the control and two psychiatric groups for their different shapes as well as different skewness and kurtosis values. The subplots (d) to (i) illustrate the pairwise comparisons of the *p*.*d*.*f*.s of the BPD and SCZ subtypes. It also strongly suggests the similarity between the subtypes BPD 0, SCZ 0, and SCZ 2, which may indicate the occurrence of mislabeled types.

On the other hand, the ranges of the SNP expression levels of the different *p*.*d*.*f*.s of the subtypes fall in different intervals on the *fSOM* plane. For example, the range of the SNP expressions of *p*.*d*.*f*.*s* of the control 0 and control 1 fall in [20, 70] and [28,70] respectively. It illustrates the good sensitivity of *fSOM* learning in uncovering the latent data characteristics of each group in a low-dimensional space.

#### 4.1 DBSCAN *pMAP* clustering and relabeling

DBSCAN prepares itself as a good candidate for *pMAP* clustering to screen mislabeled samples. The results of DBSCAN clustering demonstrate that the originally mislabeled samples sharing the same ground truth labels will be more highly likely to fall into the same cluster because of the proximity of their *pMAPs*. For example, a BPD 0 sample will be clustered into the same cluster as an SCZ 0 or SCZ 2 sample because their *pMAPs* share good similarities. The clustering result strongly suggests the possible mislabeling between them though they have similar psychiatric symptoms according to the current BPD and SCZ categorization standards [23]. Figure 5 shows the DBSCAN clusters the *pMAPs* of the three groups as 5 subclusters, where the control samples form the two subclusters are separated well from those of the BPD and SCZ samples.

**Fig 5.**
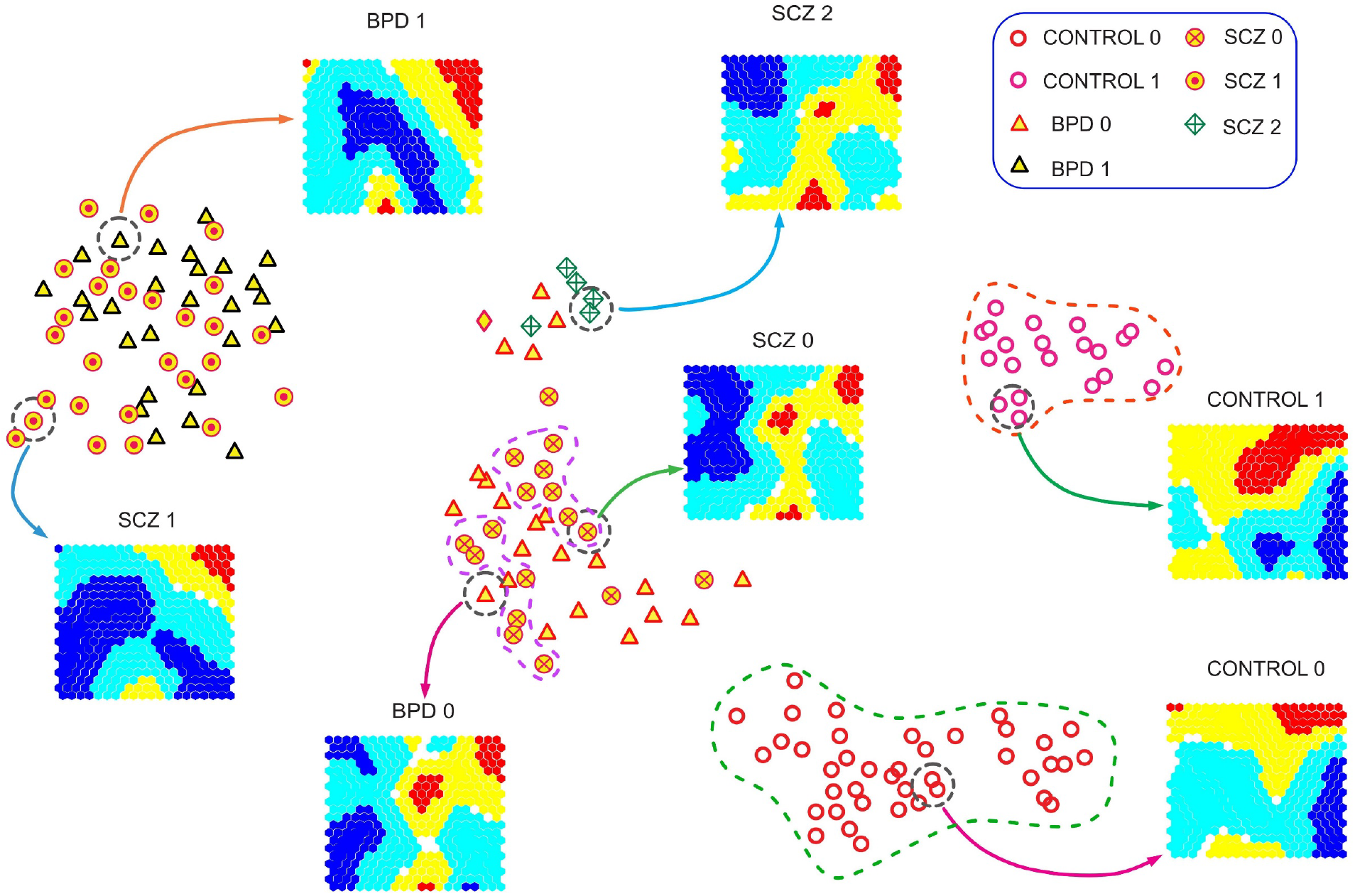
The structure of the *pMAP* clustering. The *pMAP* clustering result consists of 5 subclusters under DBSCAN: the control group partitioned as the control 0 and 1 subclusters is clearly separated from the mixed BPD and SCZ groups consisting of three subclusters. Different colors indicate different scales of the values in the *pMAP*, i.e., red and yellow symbolize the largest numerical values, and dark blue indicates the corresponding numerical values close to zero. The BPD 0, SCZ 0, and SCZ 2 samples, which are clustered in the two subclusters for their similar *pMAPs*, form a new psychiatric group BPD*. So are the clustered BPD 1 and SCZ 1 samples that generate another psychiatric group SCZ*.

Since the samples with different labels falling in the same cluster may indicate a possible mislabeling issue, we relabel them by following the rebelling rule provided in Section 2.1 to form new psychiatric groups/classes. Since the BPD 0, SCZ 0, and SCZ 2 samples are clustered in the two close subclusters and BPD 1 and SCZ 1 samples are clustered as a relatively independent subcluster, we relabel them as BPD* and SCZ* separately to reflect the ground truth better, i.e., the cluster consisting of BPD 1 and SCZ 1 samples from the BPD* group and the clusters consisting of BPD 0, SCZ 0, and SCZ 2 generate the SCZ* group. Such a clustering-based relabeling mechanism can provide a more direct and effective mislabeling information correction. Although it is possible to further exploit the statuses of each sample in DBSCAN clustering to make more refined relabeling, it may need more human intervene besides algorithmic efforts for those outlier (noise) points in each cluster.

#### 4.2 The devolution paths of psychiatric states via relative entropy analysis

##### Devolution path and intrinsic transfer

To further examine the possible pathological devolution path, we conduct a novel relative entropy analysis for the *pMAPs* after the relabeling procedure. The devolution path refers to the generic devolution process from a normal psychiatric state to dysregulated psychiatric states such as bipolar disorders or schizophrenia states. Similarly, we call the change between two subtypes of dysregulated psychiatric states an internal transfer. The reason for us to conduct such post analysis is that it cannot only unveil new knowledge about devolution paths of different psychiatric states from the normal state but also validate the correctness and effectiveness of the relabeling from the mislabel correction procedure.

The devolution path can be inferred by calculating the relative entropy, i.e., the K-L divergence of the different psychiatric states. Unlike the traditional symmetric distances, the non-symmetry of the K-L divergence provides a good measure to evaluate the devolution distance between the two psychiatric states. It is almost theoretically impossible to achieve it using the original SNP data because the probability distribution of high-dimensional SNP data is unknown. However, we can define the K-L divergence by exploiting the *pMAPs* of each psychiatric group to estimate their relative entropies.

##### K-L divergence between two psychiatric groups

Given high-dimensional datasets 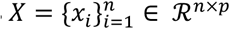 and 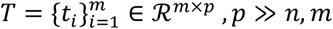 representing two different psychiatric sample groups, the K-L divergence between them is defined as,

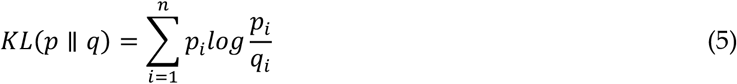

where 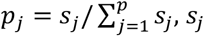 is the *j*^*th*^ singular value of the prototype data of *X Z*_*X*_ = *F*_*fsom*_(*X*) ∈ *ℛ*^*n*×*k*^ under *fSOM* learning and 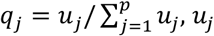 is the *j*^*th*^ singular value of *Z*_*T*_ = *F*_*fsom*_(*T*) ∈ ℛ^*m*×*k*^.

Figure 6 illustrates the K-L divergences from the two control subgroups to the other two dysregulated psychiatric states. The results seem to echo our previous relabeling result as well as provide more insights for possible devolution paths. Figure 6 (a) shows that the control 1 subgroup is closer to BPD 1 in terms of KL-divergence than control 0. It suggests that the control 1 state tends to go devolution to the BPD 1 state more likely compared to the control 0 state. That both control 0 and control 1 have almost the same K-L divergence to the SCZ 0 suggests they have the same likelihood to devote to the ‘SCZ 0’ state. Similarly, the ‘control 1’ state seems to devote to the SCZ 2 with a higher likelihood than the ‘control 0’ state. Figure 6 (b) illustrates that the. The ‘BPD 1’ state is least likely to conduct an internal transfer to the SCZ 2 compared to other dysregulated psychiatric states.

**Fig 6.**
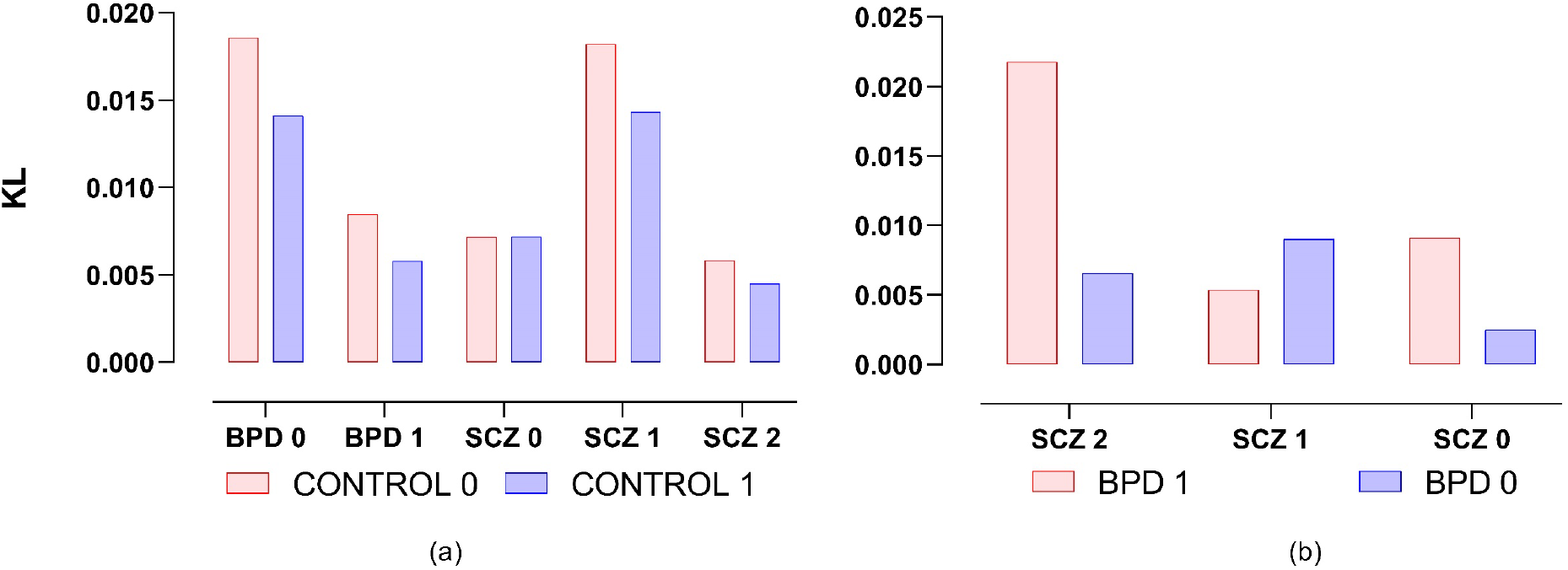
The K-L divergence analysis between the different psychiatric groups based on *pMAPs*. The subfigure (a) shows the K-L divergences of the controls 0 and 1 with respect to BPD 0, 1, and SCZ 0,1,2 respectively. The subfigure (b) describes the K-L divergence relationships between BPD 0, 1, and SCZ 0,1,2. It suggests the possible devolution paths and internal transfers between different psychiatric states.

Integrating with the previous relabeling results, we have the following interesting devolution path information. The control groups have a shorter devolution path genetically to the BPD* group for their relatively less K-L divergence values. On the other hand, a longer devolution path can exist from the control to the SCZ* group because they have a larger K-L divergence. Figure 7 illustrates the possible devolution paths of the control groups to the BPD* and SCZ* respectively according to their K-L divergences as well as internal transfers between the disease states, where the K-L divergence value is marked for each path or internal transfer. Classic psychiatric studies seem to support the devolution path because a subject can start from a normal psychiatric state to a more dysregulated, complicated, or unstable state in a gradual or abrupt manner. Moreover, previous studies also support the finding because it was reported that bipolar disorder can be a transition state between the normal and schizophrenia states [38].

**Fig 7.**
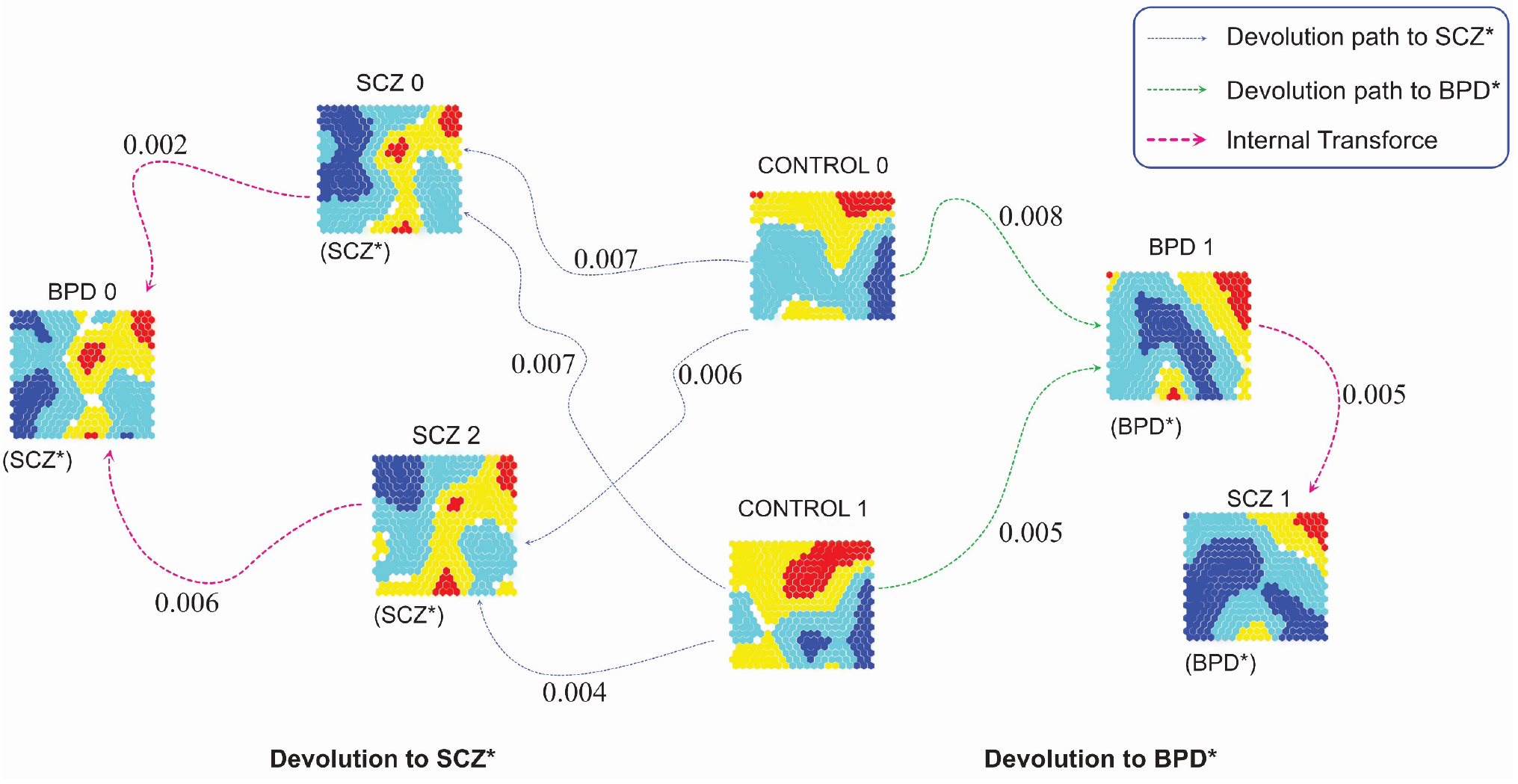
The possible devolution paths of the control groups to the BPD* and SCZ* according to their pMAP patterns. The pMAPs of the BPD* and SCZ* groups have more complicated patterns than those of controls. It indicates that the BPD* is an intermediate psychiatric state between the SCZ* and control.

## 5 The comparisons of psychiatric map diagnosis with peer methods

We conduct control, BPD, and SCZ diagnosis with the relabeled data to validate the correctness and effectiveness of the relabeling. The effective relabeling should lead to good improvements in diagnostic accuracy and poor or mediocre diagnosis performance may suggest the relabeling does not do a good correction for the noisy labels. In other words, if the ground truth labels are assumed as the relabeled ones, it should show improvements in comparison with the previous labels if the relabeling is correct and solid enough under a reproducible machine learning model, which is selected as multi-SVM in this study.

Since the traditional classification measures are neither efficient nor interpretable in assessing different machine learning models’ performance, we extend the proposed diagnostic index (d-index) measure under binary classification to provide a more explainable and sensitive learning performance evaluation [39-40]. This is because the traditional classification measure assessment may only reflect one aspect of classification performance. As a result, it is inconvenient to compare many classification measures for different machine learning model performances on different datasets in a more explainable approach. The d-index definition of the binary classification can be found in the following section and more d-index information can be found in [40].

### Diagnostic index (d-index)

Given a prediction function 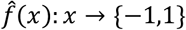 constructed from training data 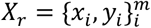 under a machine learning model *Θ*, where each sample *x*_*i*_ ∈ *ℛ*^*p*^ and its label *y*_*i*_ ∈{−1,1}, *i* = 1,2, ⋯ *m*, d-index evaluates the performance of 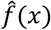 in predicting the class of test data 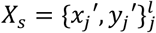. It is defined as

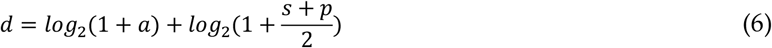

where *a, s* and *p* represent the corresponding accuracy, sensitivity, and specificity in diagnosing test data respectively. The larger the d-index value, the better the predictability of 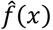, *i*.*e*., the better learning performance achieved by the machine learning model *Θ*. The maximum value of d-index is 2 where classification has the perfect results. The minimum value of the d-index is 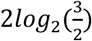 if there is no underfitting [40].

Figure 8 compares the d-index values and misclassification rates before and after relabeling in detecting control, BPD, and SCZ, under four machine learning models: SVM, random forests (RF), extremely randomized trees (ET), and deep neural networks (DNN), where training and test datasets have 80% and 20% samples of the total samples respectively [15, 30, 31]. We employ nSVA to select *p*% (p=10, 20, …100) top-ranked features to observe how the relabeling results impact those datasets under different percentages of top-feature selection. The d-index values of the diagnoses after relabeling are much higher than those before relabeling for all datasets across all four models under the nSVA feature selection. Correspondingly, the error rates of learning performance decreases significantly for those relabeled data. It strongly suggests the correctness and effectiveness of the relabeling. Furthermore, all the models have low misclassification rates after relabeling and SVM had the lowest ones, which indicates the strong reproducibility of the proposed psychiatric map diagnosis.

**Fig 8.**
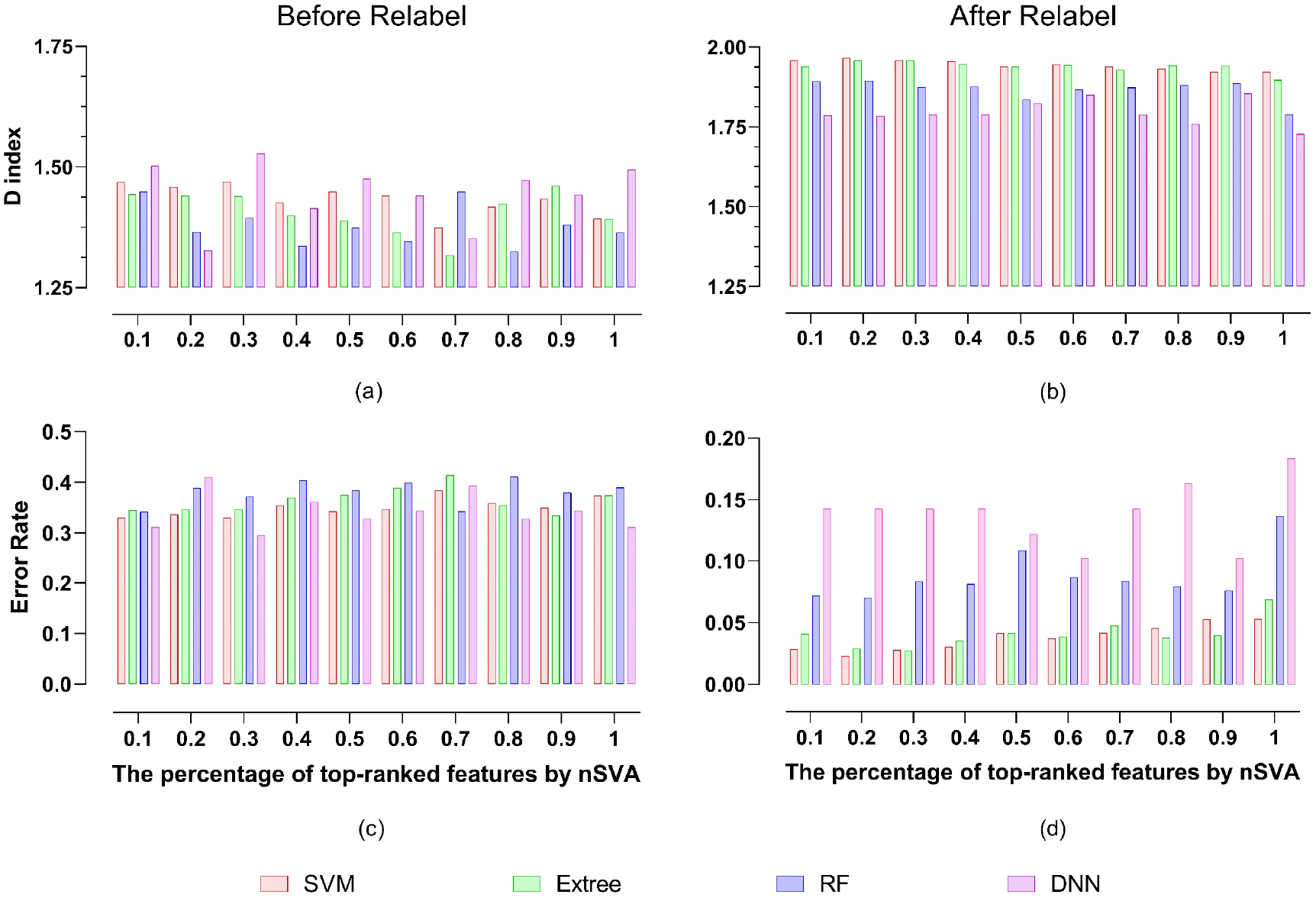
The comparisons of control, BPD, and SCZ diagnoses before and after relabeling under nSVA feature selection under four machine learning models.

Figure 9 compares the precision, recall, and F1 values under multi-class SVM as well as entropy values before and after relabeling for different psychiatric groups under different percentages of features selected by nSVA. The precision, recall, and F1 values are consistent with the d-index values well. Interestingly, we have found that the original BPD and SCZ groups only show they have relatively smaller entropy values than that of the control group and the relationship of the entropies of the BPD and SCZ groups looks ‘random’. However, the entropies of the relabeled subgroups demonstrate more regular patterns: the relabeled BPD* subgroup regularly has the smallest entropies, and SCZ* has the second-smallest entropies. The results somewhat validate the correctness of relabeling besides suggesting that the psychiatric disorder samples may have more special SNP patterns than those of the normal ones.

**Fig 9.**
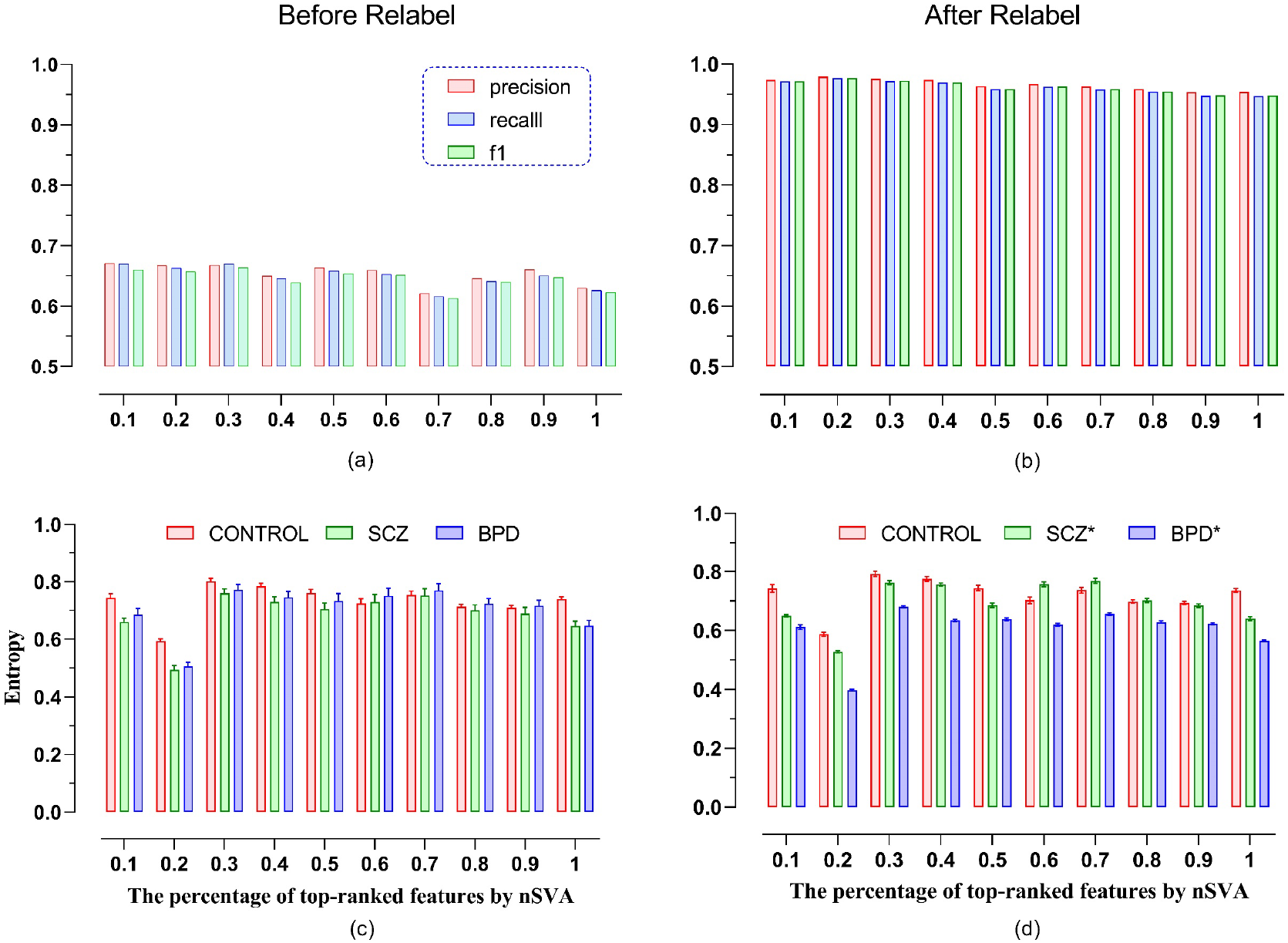
Classification metric and entropy comparisons before and after relabeling under different percentages of top-ranked features by nSVA. The subfigures (a) and (b) compares the values of recall precision, F1-score before and after relabeling before and after relabeling under multi-class SVM. The subfigures (c) and (d) illustrate the differences of entropies of control, SCZ, and BPD.

#### 5.1 Comparison with state-of-the-art machine learning methods

We compare our results with those of state-of-the-art ML and deep learning models to further validate the effectiveness of the proposed *pMAP* diagnosis. The comparison methods include one-shot learning, convolutional neural networks (CNN), residual neural networks (ResNet), long short-term memory (LSTM), Transformer, and generative adversarial networks (GAN), as well as classic support vector machines (SVM) [41-46]. The comparison results can alternatively answer the query: ‘what will happen to the deep learning and ML models under mislabeled high-dimensional data?’

We briefly describe the deep learning models for the convenience of description. CNN is characterized by a partially connected layer structure and different layers have different functionalities such as convolutional, and max/average pooling. It demonstrates powerful learning capabilities, especially for image data besides decent feature extraction. ResNet is an enhanced CNN using residual learning techniques to tackle the challenges of gradient disappearance and explosion in CNN learning. It demonstrates advantages over CNN in handling big and more complicated data. LSTM, which is widely employed in time-series data analysis, overcomes the weakness of general recurrent neural networks (RNN) in handling long-time information dependence by employing LSTM cells that consist of three different gates. GAN employs two different neural networks: a generator and discriminator (e.g., CNN and LSTM) to contest with each other to accomplish learning. GAN stops at the point when the discriminator was completed ‘confused’ by the learning results from the generator. We apply the DNN model to the data obtained from GAN. In addition, Transformer is a special feedforward neural network taking advantage of the self-attention mechanism in topology and learning. It improves the parallelism of the model and decreases its reliance on long-term memory. One-short learning aims to handle the data scarcity issue in deep learning, i.e., input data itself is small enough to satisfy the training demand for the number of observations in training. It creates models that can accurately identify test samples with a limited quantity of training data. More details about the models can be found in the literature [41,47].

Figure 10 compares the proposed *pMAP* diagnosis with the state-of-the-art deep learning models as well as SVM which is a representative of the classic learning method, under different levels of feature selection by nSVA. It is obvious that the *pMAP* diagnosis demonstrates its superiority to the rest of the methods no matter in learning effectiveness or stability. It suggests that all deep learning models show quite poor performance on the original data. For example, LSTM obviously fails the whole learning process by encountering overfitting because of its very low d-index value [40].

**Fig 10.**
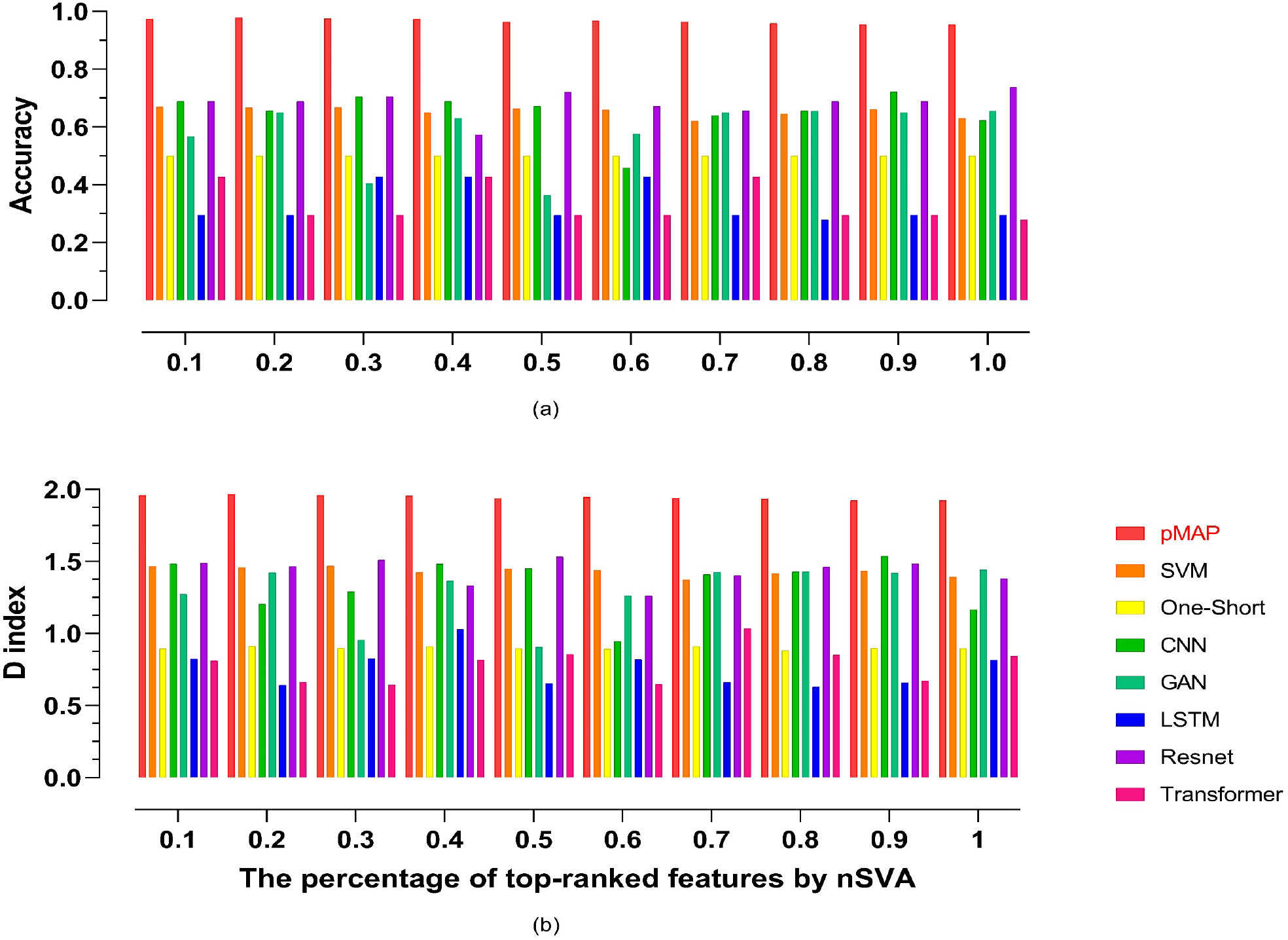
The comparisons of the proposed *pMAP* diagnosis with its peer methods: SVM, one-short learning, CNN, GAN, LSTM, ResNet, and Transformer. The *pMAP* diagnosis demonstrates stably leading performance compared to its peers under different levels of feature selections. Almost all deep learning models show poor performance for the original data. Both one-short learning and LSTM encounter underfitting.

As we pointed out that deep learning may not work well for mislabeled high-dimensional data learning. The two reasons can interpret the poor performance from the deep learning models. The first is the issue of the data scarcity issue, i.e., the high-dimensional data has a small number of samples that prevents it from taking advantage of the powerful learning capabilities of the deep learning models. The second, which can be more important, is the problem itself is a mislabeled learning problem, but there are no deep learning techniques available to handle mislabeled high-dimensional data. This also can explain why one-short learning encounters poor performance because of underfitting.

On the other hand, why the *pMAP* diagnosis leads all the other methods lies in that it is a specifically designed algorithm for mislabeled learning. It exploits *fSOM* learning to gain the *pMAP* for each SNP observation and density clustering to seek the similarities between the *pMAPs*. It takes advantage of the DBSCAN clustering results of the prototypes of the original observations to relabel data to decrease mislabeling information at the most level. Finally, the kernel method SVM is employed to conduct psychiatry prediction by exploiting its reproducibility and efficiency. Therefore, the *pMAP* diagnosis is more effective, efficient, and robust in handling the mislabeling psychiatry learning problem than its possible peer methods.

## 6 Discussion and conclusion

We propose a novel mislabeled learning algorithm for high-dimensional data to provide a technique to overcome the long-time misdiagnosis between BPD and SCZ in psychiatry from an AI perspective. To the best of our knowledge, mislabeled learning for high-dimensional data is a rarely investigated but essentially important and challenging problem in modern AI and data science. With the surge of big data and AI, more and more mislabeled learning problems need serious investigations according to their ‘data background’. Simply assuming the label information is correct would cause ML to encounter mediocre or poor performance and produce a serious misdiagnosis in many AI-driven disease diagnoses (e.g., mental disorder detection) and present a hurdle in AI health and related interacting fields.

The proposed *pMAP* diagnosis employs feature self-organizing map (*fSOM*) learning to tackle mislabeled learning for high-dimensional data successfully. It generates a *pMAP* that is essentially the 2D characteristic image for each high-dimensional observation. The *pMAPs* discover and unveil new knowledge for input data besides disclosing their latent data distributions. They also identify different unknown hidden subtypes for each group. For example, it finds that there are two subtypes in the control group, 2 subtypes in the BPD, and 3 subtypes in the SCZ group. We employ a novel relabeling technique to correct label information according to DBSCAN clustering for the 2D characteristic images of input data, before employing multi-SVM for high-accuracy psychiatric diagnosis. It provides a more explainable and implementable technique without human intervene to overcome the long-time misdiagnosis issue in psychiatry from AI perspective and sheds new light on mislabeled learning for high-dimensional data.

Furthermore, the devolution path of psychiatric states via relative entropy analysis provides insights into existing pathological psychiatry. The novel devolution path analysis unveils latent internal transfer and devolution road maps between different subtypes of the control, BPD, and SCZ groups, which have been rarely investigated in the existing psychiatry studies and bioinformatics research. It will inspire more future studies on this topic via similar devolution path analysis ways. However, it still may need more data and experiments or genetic findings to validate its effectiveness furthermore.

Although this method is designed to tackle mislabeled learning for high-dimensional data, it can be theoretically applied or extended to more general mislabeled learning because its essential components such as prototype learning, DBSCAN clustering, multiclass SVM can essentially apply to any kind of data. However, it needs to point out that *fSOM* assumes input data should be high-dimensional data rather than general data. It may not get desirable results by just applying it to other data because the variable space may not be that meaningful in the low-dimensional embedding. It is also noted that *fSOM* may need a huge computing demand in generating *pMAPs* especially because of the high dimensionality of input data. We have spent about 2 weeks completing *fSOM* learning on a 20×20 *fSOM* plane to generate 203 *pMAPs* on an Intel Xeon E5-2620 machine under OS Ubuntu 20.04 LST (Focal Fossa) with RAM 128Gb and CPU speed 2.1Ghz. We are seeking to implement *fSOM* via an FPGA approach to tackle the high computing demands [48].

In addition, there are quite a few aspects to be improved in the proposed methods. For example, it is desirable to design more customized data-driven kernels in multiclass SVM rather than rely on some standard nonlinear kernels (e.g., Gaussian kernel). Furthermore, we are interested in exploring to build new interpretable machine learning models by employing new techniques such as hierarchical level *fSOM* learning structure or FPGA speedup to decrease its complexity for the sake of scalability and adaptability, especially for large-scale high-dimensional data. In addition, it can be possible to substitute the existing DBSCAN by DBSCAN++, HDBSCAN, OPTICS or other variants such as OPTICS to handle large data and get better *pMAP* clustering performance before relabeling [49-51]. Last but the least, we are also interested in continuing our investigations in AI-based devolution path in psychiatry.

## Data Availability

https://github.com/AidenLee1994/Psychiatric-map-diagnosis-in-mental-disorders/data

https://github.com/AidenLee1994/Psychiatric-map-diagnosis-in-mental-disorders

## Data availability

The data of this work is publicly available at: https://github.com/AidenLee1994/Psychiatric-map-diagnosis-in-mental-disorders/data

## Code availability

The code of this work is publicly available at: https://github.com/AidenLee1994/Psychiatric-map-diagnosis-in-mental-disorders

## Acknowledgment

This work was supported in part by the National Science Foundation of China under Grant 61572367, Grant 61573017,and McCollum endowed chair startup fund.

## Authors’ contributions

Han designed the methods and algorithms. Li, Liu, and Han analyzed the data. Han and Liu wrote the manuscript. Han finalized the paper.

## Reference

[1] Song et al (2022) Learning from Noisy Labels with Deep Neural Networks: A Survey, IEEE transactions on neural networks and learning systems, 3152527, 2022

[2] Ayano, G., Demelash, S., yohannes, Z. et al.. Misdiagnosis, detection rate, and associated factors of severe psychiatric disorders in specialized psychiatry centers in Ethiopia. Ann Gen Psychiatry 20, 10 (2021). https://doi.org/10.1186/s12991-021-00333-7

[3] H. Shen, L. Zhang, C. Xu, J. Zhu, M. Chen, and Y. Fang, “Analysis of Misdiagnosis of Bipolar Disorder in An Outpatient Setting,” Shanghai archives of psychiatry, vol. 30, no. 2, pp. 93–101, 2018, doi: 10.11919/j.issn.1002-0829.217080.

[4] Natarajan, N, Dhillon, I, and Ravikumar, P, Tewari, A: Learning with noisy labels, NIPS 2013

[5] Lee, K, He, X, Zhang, Lei, Yang, L: CleanNet: Transfer Learning for Scalable Image Classifier Training with Label Noise, CVPR 2018.

[6] Eric Arazo, Diego Ortego, Paul Albert, Noel E. O’Connor, and Kevin McGuinness. Unsupervised label noise modeling and loss correction. ICML, 312–321, 2019.

[7] Berthelot et al: Mixmatch: A holistic approach to semi-supervised learning. NeurIPS, 2019.

[8] Han et al Co-teaching: Robust training of deep neural networks with extremely noisy labels. In NeurIPS, pp. 8536–8546, 2018.

[9] Witt, S., Streit, F., Jungkunz, M. et al.. Genome-wide association study of borderline personality disorder reveals genetic overlap with bipolar disorder, major depression and schizophrenia. Transl Psychiatry 7, e1155 (2017). https://doi.org/10.1038/tp.2017.11

[10] Karege et al. “Genetic overlap between schizophrenia and bipolar disorder: a study with AKT1 gene variants and clinical phenotypes.” Schizophrenia research 135.1-3 (2012): 8-14.

[11] Antonio et al. “Effectiveness, core elements, and moderators of response of cognitive remediation for schizophrenia: a systematic review and meta-analysis of randomized clinical trials.” JAMA psychiatry 78.8 (2021): 848–858.

[12] Sahu et al. Integrative network analysis identifies differential regulation of neuroimmune system in Schizophrenia and Bipolar disorder. Brain, Behavior, & Immunity-Health 2 (2020): 100023.

[13] Li et al. Altered DNA methylation of the Alu Y subfamily in schizophrenia and bipolar disorder. Epigenomics 11.6 (2019): 581–586.

[14] Ellis et al. Transcriptome analysis of cortical tissue reveals shared sets of downregulated genes in autism and schizophrenia.” Translational psychiatry 6.5 (2016): e817–e817.

[15] Liu, W, Li, D, and Han, H Manifold learning analysis for allele-skewed DNA modification SNPs for psychiatric disorders. IEEE Access 8 (2020): 33023–33038.

[16] Birur et al. cBrain structure, function, and neurochemistry in schizophrenia and bipolar disorder—a systematic review of the magnetic resonance neuroimaging literature. NPJ schizophrenia 3.1 (2017): 1–15.

[17] Oh et al. Identifying schizophrenia using structural MRI with a deep learning algorithm. Frontiers in psychiatry 11 (2020): 16.

[18] Dubovsky et al. Psychotic depression: Diagnosis, differential diagnosis, and treatment.” Psychotherapy and Psychosomatics 90.3 (2021): 160–177.

[19] Ayano et al. Misdiagnosis, detection rate, and associated factors of severe psychiatric disorders in specialized psychiatry centers in Ethiopia. Annals of general psychiatry 20.1 (2021): 1–10.

[20] Kohonen, T. Self-organizing neural projections. Neural networks 19.6-7 (2006): 723-733.

[21] Han, H. A novel feature selection for RNA-seq analysis, Computational biology and chemistry 71 (2017): 245–257.

[22] Bansal, M, Sharma, R, Kathuria, M: A Systematic Review on Data Scarcity Problem in Deep Learning: Solution and Applications.” ACM Computing Surveys (CSUR) (2021).

[23] Geschwind, D, and Flint, J: Genetics and genomics of psychiatric disease, Science 349.6255 (2015): 1489–1494.

[24] Jiang, B, Ding, C, and Luo, B (2018). Robust data representation using locally linear embedding guided PCA. Neurocomputing, 275:523–532

[25] Han et al. Enhance Explainability of Manifold Learning. Neurocomputing 500:877–895 2022.

[26] Der Maaten, L, Hinton, G (2008) Visualizing High-Dimensional Data Using t-SNE. Journal of Machine Learning Research, 9:2579–2605.

[27] Schubert et al. DBSCAN revisited, revisited: why and how you should (still) use DBSCAN, ACM Transactions on Database Systems (TODS) 42.3 (2017): 1–21.

[28] Ester et al. A density-based algorithm for discovering clusters in large spatial databases with noise, KDD 96:34. 1996.

[29] Crammer, K, Singer Y (2001) On the Algorithmic Implementation of Multiclass Kernel-based Vector Machines, JMLR, 265–292

[30] Geurts, P, Ernst, D and Wehenkel, L: Extremely randomized trees. Machine learning 63.1 (2006): 3–42.

[31] Wu, and Weng, Probability estimates for multi-class classification by pairwise coupling, JMLR 5:975–1005, 2004

[32] Bergstra, J. and Bengio, Y., Random search for hyper-parameter optimization, The Journal of Machine Learning Research (2012)

[33] Becht et al (2019): UMAP: Uniform Manifold Approximation and Projection for Dimension Reduction, Nature Biotech, 37: 38–44

[34] Gagliano et al. Allele-skewed DNA modification in the brain: relevance to a schizophrenia GWAS, The American Journal of Human Genetics 98.5 (2016): 956–962.

[35] Meaburn, E, Schalkwyk, L, and Mill, J. Allele-specific methylation in the human genome: implications for genetic studies of complex disease. Epigenetics 5.7 (2010): 578–582.

[36] Russell and Norvig (2020). Artificial intelligence a modern approach, 4th ed. Prentice hall, 2022

[37] Chen, Yen-Chi. A tutorial on kernel density estimation and recent advances. Biostatistics & Epidemiology 1.1 (2017): 161–187.

[38] Santa-Cruz et al. Association between trace elements in serum from bipolar disorder and schizophrenia patients considering treatment effects. Journal of Trace Elements in Medicine and Biology 59 (2020): 126467.

[39] Han, H, Men, K. How does normalization impact RNA-seq disease diagnosis? Journal of biomedical informatics 85 (2018): 80–92.

[40] Han et al. Interpretable Machine Learning Assessment. Available at SSRN: https://ssrn.com/abstract=4146556

[41] Vinyals et al. Matching networks for one shot learning, Advances in neural information processing systems 29 (2016).

[42] Gu et al. Recent advances in convolutional neural networks, Pattern Recognition 77 (2018): 354–377.

[43] Xie et al. Aggregated residual transformations for deep neural networks, Proceedings of the IEEE conference on computer vision and pattern recognition (CVPR). 2017.

[44] Hochreiter and Schmidhuber, Long short-term memory. Neural computation 9.8 (1997): 1735–1780.

[45] Han et al. “Transformer in transformer.” Advances in Neural Information Processing Systems 34 (2021).

[46] Creswell et al. Generative adversarial networks: An overview. IEEE Signal Processing Magazine 35.1 (2018): 53–65.

[47] Li, F, Fergus, R, and Perona, P, One-shot learning of object categories, IEEE transactions on pattern analysis and machine intelligence 28.4 (2006): 594–611.

[48] Kurdthongmee et al. A novel hardware-oriented Kohonen SOM image compression algorithm and its FPGA implementation, Journal of Systems Architecture 54.10 (2008): 983–994.

[49] Campello, R, Moulavi, D, Zimek, A, Sander, J (2015). “Hierarchical Density Estimates for Data Clustering, Visualization, and Outlier Detection”. ACM Transactions on Knowledge Discovery from Data. 10 (1): 1–51

[50] Ankerst, M, Breunig, K, Kriegel, H, and Sander, J. OPTICS: ordering points to identify the clustering structure. ACM SIGMOD 18:49-60, 1999

[51] Jang and Jiang (2019) DBSCAN++: Towards fast and scalable density clustering ICML, 2019

